# Comparing neural language models for medical concept representation and patient trajectory prediction

**DOI:** 10.1101/2023.06.01.23290824

**Authors:** Alban Bornet, Dimitrios Proios, Anthony Yazdani, Fernando Jaume Santero, Guy Haller, Edward Choi, Douglas Teodoro

## Abstract

Effective representation of medical concepts is crucial for secondary analyses of electronic health records. Neural language models have shown promise in automatically deriving medical concept representations from clinical data. However, the comparative performance of different language models for creating these empirical representations, and the extent to which they encode medical semantics, has not been extensively studied. This study aims to address this gap by evaluating the effectiveness of three popular language models – word2vec, fastText, and GloVe – in creating medical concept embeddings that capture their semantic meaning. By using a large dataset of digital health records, we created patient trajectories and used them to train the language models. We then assessed the ability of the learned embeddings to encode semantics through an explicit comparison with biomedical terminologies, and implicitly by predicting patient outcomes and trajectories with different levels of available information. Our qualitative analysis shows that empirical clusters of embeddings learned by fastText exhibit the highest similarity with theoretical clustering patterns obtained from biomedical terminologies, with a similarity score between empirical and theoretical clusters of 0.88, 0.80, and 0.92 for diagnosis, procedure, and medication codes, respectively. Conversely, for outcome prediction, word2vec and GloVe tend to outperform fastText, with the former achieving AUROC as high as 0.78, 0.62, and 0.85 for length-of-stay, readmission, and mortality prediction, respectively. In predicting medical codes in patient trajectories, GloVe achieves the highest performance for diagnosis and medication codes (AUPRC of 0.45 and of 0.81, respectively) at the highest level of the semantic hierarchy, while fastText outperforms the other models for procedure codes (AUPRC of 0.66). Our study demonstrates that subword information is crucial for learning medical concept representations, but global embedding vectors are better suited for more high-level downstream tasks, such as trajectory prediction. Thus, these models can be harnessed to learn representations that convey clinical meaning, and our insights highlight the potential of using machine learning techniques to semantically encode medical data.

## 1. Introduction

With the widespread adoption of electronic health record (EHR) systems in healthcare institutions, medical research has increasingly focused on the secondary use of clinical data for improving healthcare management and cost [1], classifying diseases [2, 3], handling complex medical data [4], and advancing medical research [5]. Moreover, events from discharge summaries and clinical notes can be used to identify adverse drug events [6–9] and improving patient quality outcomes at the individual [10–12] and population [13] levels. This growing interest extends to phenotype extraction for clinical and genetic research [14, 15], patient cohort identification [16–18], clinical outcome and patient trajectory prediction [19–22], clinical decision support [23–26], and decision making [27, 28]. However, even though big data analytics in healthcare offers promising insights for improving outcomes and reducing costs, it faces challenges due to the complexity and heterogeneity of healthcare data [29–32].

Classic approaches to abstract patient data and create homogeneous representations for secondary analyses are based on data mappings [33–36], in which raw data are encoded using concepts from biomedical knowledge organization systems [37], such as the International Classification of Diseases (ICD) [38], the Anatomical Therapeutic Chemical (ATC) [39] classification systems, and the Systematized Nomenclature in Medicine – Clinical Terms (SNOMED-CT) ontology [40]. Despite the benefits of these approaches for representing knowledge in EHRs and the facilitated semantic interoperability, there are limitations and challenges that need to be considered. First, large human efforts are required to curate and annotate data, a resource that is often not at one’s disposal in hospitals [41–43]. Additionally, as data and information evolve, the lack of readily available and up-to-date formal representations might hinder their application to the full extent of EHR data [44]. Lastly, the resulting representations of medical concepts are highly dimensional, leading to sparse and computationally inefficient data structures (e.g., SNOMED-CT alone has more than 300’000 concepts).

In recent years, as a complementary alternative to fully semantic encoding, data-driven methods for EHR concept representation based on deep learning were proposed [45–48]. In contrast to knowledge-based approaches, deep learning algorithms learn representations of patients and clinical concepts automatically and directly from the data, with minimal pre-processing. The learnt representations are dense, low-dimensional vectors that can be used in many downstream tasks. This approach, known as medical concept embedding, has already achieved promising results (for reviews: [49–52]). For example, convolutional, long short-term memory, as well as attention-based neural networks were trained with patient data to perform various patient trajectory prediction tasks [53–57]. Moreover, similar neural network architectures were used for automated phenotyping, by building representations of structured [58–61], unstructured [62–65] EHR data, or both [66–69]. As a recent use-case scenario, deep learning methods were applied to COVID-19-related EHR data for epidemiological prediction [70], automatic diagnosis [71], drug repurposing [72–75], or mortality risk assessment [76–78].

A key aspect of developing accurate patient and medical concept embeddings lies in abstracting and modeling the relevant data features. In that respect, neural language models can process EHRs, not only for clinical notes, but also by considering events in the patient trajectory as medical concept tokens, and entire trajectories as a sequence of tokens, similar to word sentences [79–84]. Medical concepts relate to each other, either with causal relationships, e.g., diagnosis and medication, or through similar meanings, e.g., related, or synonymous diagnoses or compounds with similar effects or indications. Analogous to text, syntactic and semantic relationships among EHR concepts can thus be learned by neural language models: leveraging this inherent data structure was shown to improve representations of medical concepts and patients [48, 85]. For example, attention-based language models were trained with sequences of clinical events and produced representations which improved performance for disease prediction [54] as well as for length-of-stay, readmission, and mortality prediction [86, 87].

Several studies compared the advantages and shortcomings of popular neural language models, such as word2vec [88], fastText [89, 90] and Global Vectors for Word Representation (GloVe) [91], to create word embeddings when applied to clinical notes [92–95]. Still, no study has yet performed a comprehensive comparison for medical concept code representations. In this work, we aim to assess the performance of neural language models to generate embeddings from patients’ stays at the hospital expressed as sequences of healthcare related event codes. In previous work, using the Medical Information Mart for Intensive Care (MIMIC) IV dataset [96], we showed that embeddings produced by word2vec provide useful representations of medical concept codes [83]. In this work, we extend this study by comparing the quality of embeddings produced by word2vec, fastText and GloVe. To do so, we create patient trajectories as sequences of administrative, demographic, and clinical events using the MIMIC-IV dataset. Then, the different neural language models are trained to learn medical concept representations from these trajectories. Finally, we qualitatively evaluate the extracted representations using a clustering task (intrinsic evaluation) and quantitatively using binary and categorical trajectory prediction tasks (extrinsic evaluation). The code that we used to build patient trajectories, train the models, and evaluate them, is available on our repository^1^.

The contribution of this work can be summarized as follows:

1. We compare popular neural language model architectures – word2vec, fastText and GloVe – trained on patient trajectories created from sequences of medical codes and show that these language models can produce data-driven embeddings that capture the semantic meaning of medical concepts, as defined by biomedical terminologies.
2. We evaluate the alignment between medical concept embeddings and biomedical terminologies and, using a clustering algorithm, we show that fastText naturally presents the highest similarity to the different hierarchical levels of the terminologies, with a cluster similarity distance between 0.80 and 0.92 for diagnosis, procedure, and medication codes.
3. We assess patient outcome prediction at different information levels and show that after 10% of the trajectory, the extracted embeddings can estimate mortality risks with performance above 0.80 AUROC. On the other hand, using the full trajectory (i.e., 100% of patient tokens), we can estimate readmission with performance of only around 0.60 AUROC.
4. Lastly, we evaluate how much intrinsic information the learned embeddings encode to infer the next events in the patient trajectory, i.e., diagnoses, procedures, and medications. We demonstrate that while high-level information can be encoded by the embeddings, with performance varying from 0.45 AUPRC for diagnosis codes to 0.81 AUPRC for medication codes, prediction performance decay exponentially with the increase of semantic granularity.

## 2. Methodology

### 2.1 MIMIC-IV dataset and pre-processing steps

We extracted 431,231 hospital stays from the 299,712 patients present in the MIMIC-IV database [96]. MIMIC-IV is a large, openly available dataset of de-identified EHRs from patients admitted to intensive care units (ICUs) or to the emergency department at Beth Israel Deaconess Medical Center in Boston between 2008 and 2019. The dataset contains comprehensive clinical data, including vital signs, laboratory results, medications, procedures, and diagnoses, as well as demographic information such as age, gender, and race. All entries in MIMIC-IV are associated with a patient and an admission identifier, and most of them are (directly or indirectly) labeled with a timestamp. This allowed us to assemble sequences of events occurring during patients’ hospital stays. Each patient may be admitted several times at the hospital, and for each sequence of events happening between admission and discharge time, we computed one sequence of events, which will be referred to as “*patient trajectory*” from now on. Each event was extracted from MIMIC-IV tables as a token which could encode a label associated with the patient demographic, administrative information, or a medical event (Figure 1). We provide a detailed description of each token category in the supplementary information, Appendix A1.

**Figure 1.**
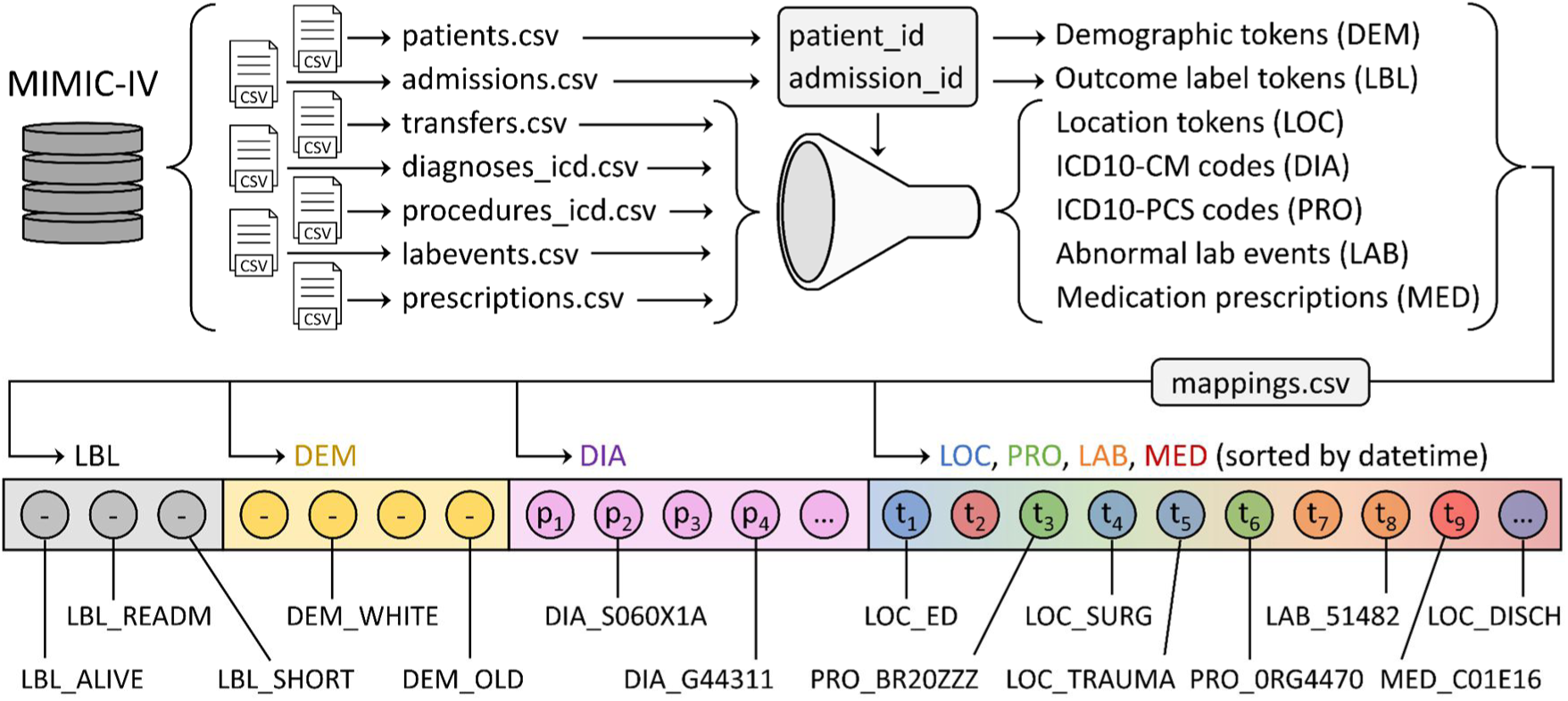
Patient trajectory sequence generation. Medical data was extracted from the MIMIC-IV database. Patient and admission ids were used to parse the data and obtain one sequence per patient trajectory. Outcome labels and demographic tokens were built using patient and admission data. Token sequences were built from the MIMIC tables, using mappings from MIMIC or custom mappings. Tokens for location, procedures, abnormal lab events and medication prescriptions were sorted by datetime. Outcomes, demographic information, and ICD10-CM codes were prepended to the sequence, because they do not have any associated datetime. ICD10-CM codes were sorted by priority.

As shown in Figure 1, patient trajectories were built by concatenating the tokens from the outcome (LBL), demographic (DEM), diagnosis (DIA), administrative (LOC), procedure (PROC), laboratory (LAB) and medication (MED) categories. The outcome and demographic tokens started each sequence, and diagnosis codes came second, as a reason for intensive care hospitalization. The remaining tokens were appended to the sequence, after being sorted altogether by their associated datetimes (Figure 1). The different token categories extracted from the MIMIC database are summarized in Table 1.

**Table 1.** Summary of the tokens used to build patient trajectory sequences. The number of tokens per sequence indicates the mean and the standard deviation, which were computed from the training dataset. Note that, in MIMIC-IV, all medications are stored as Generic Sequence Number (GSN) or National Drug Code (NDC) entries. We mapped these entries to their Anatomical Therapeutic Chemical (ATC) code equivalents, by using the work of Kury et al. [97], which queries the online RxNorm API [98] automatically. ATC codes are hierarchically arranged following the target anatomical system of medications, as well as their therapeutic, pharmacological, and chemical effects. Appendix A1 describes in detail how we built the mapping, which we make available in our repository^2^.

| Category | Content | Encoding | Token instances (#) | Tokens per sequence (#) |
| --- | --- | --- | --- | --- |
| LBL | Labels for binary classification | Custom | 6 | 3.00 ± 0.00 |
| DEM | Gender, age, race | Custom | 37 | 3.28 ± 0.45 |
| DIA | Diagnoses and reason for visits | ICD10-CM | 9774 | 11.01 ± 7.29 |
| LOC | Care unit locations | Custom | 28 | 5.22 ± 2.99 |
| PRO | Procedures | ICD10-PCS | 4243 | 1.53 ± 2.40 |
| LAB | Lab results outside normal range | Ids | 398 | 49.39 ± 127.99 |
| MED | Medication prescriptions | ATC | 1122 | 31.57 ± 41.34 |

Once all sequences were constructed, the dataset was split into training, validation and testing subsets, containing 345.3k, 43.5k, and 42.4k patient trajectory sequences, respectively. The splitting was based on patient ids (80% for training, 10% for validation, 10% for testing) and not admission ids, which explains the different number of samples for the validation and testing sets. This means that for any patient, all patient trajectories were included in the same data subset. This avoided patient information leaking from the training data to the validation and testing data.

### 2.2 Training language models

We trained three language models – word2vec [88], fastText [89, 90], and GloVe [91] – to compute medical concept embeddings for any token appearing in the patient trajectory sequences. Patient trajectory sequences were encoded by a vocabulary that assigned an integer id to any token appearing at least 5 times in the training data and a special id for the remaining “unknown” tokens. Each model mapped these ids to fixed-size float vectors, i.e., embeddings, initialized with a random floating point look-up table. We used an embedding dimensionality of 512 for all models. These embeddings were trained by optimizing the different model’s objectives, in order to provide useful representations of medical concepts (Figure 2, top). Because of the unique nature of our dataset (comprising medical concept tokens, i.e., no natural language), models were trained from scratch.

**Figure 2.**
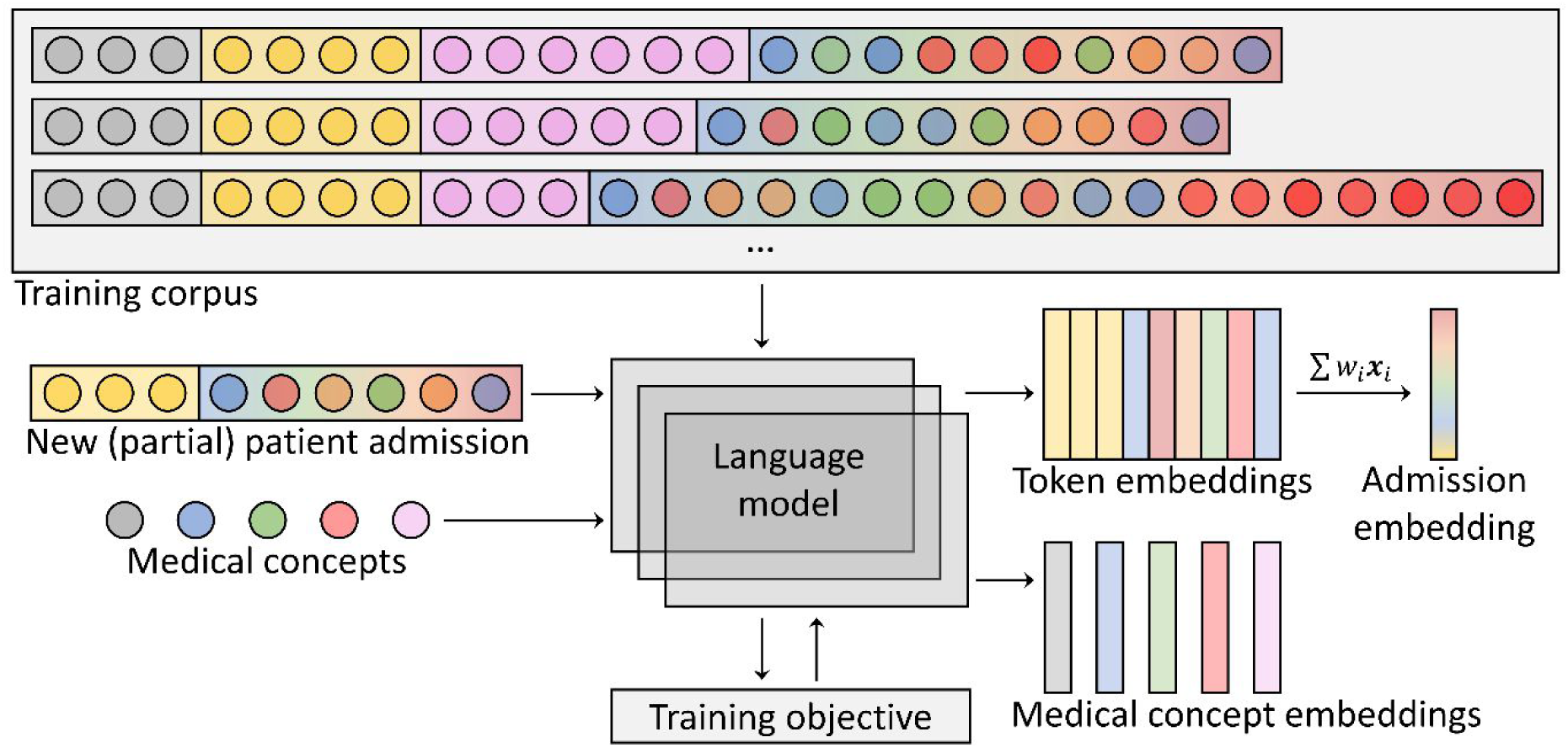
Training and evaluation of language models. Language models (word2vec, fastText or GloVe) were trained, based on their respective objectives (see sections 2.2.1 to 2.2.3 for details), using patient trajectories from the training dataset. After training, embeddings of patient trajectories or single medical concepts were queried using the hidden weights of the models. To generate a single embedding vector for a patient trajectory, token embeddings were averaged, weighting each token by the inverse of its frequency in the training dataset.

We trained all models until convergence – 100’000 training steps for word2vec and fastText, and 300’000 for GloVe – with a batch size of 4096 tokens. After training, any medical concept could be given as input to the language model to obtain a vector representation (Figure 2, bottom). To embed any patient trajectory, we computed a weighted average of the embeddings of its tokens (Figure 2, bottom-right). The weights were defined as the inverse token frequencies in the training dataset. The neural language models used in our experiments are described in sections 2.2.1 to 2.2.3.

#### 2.2.1 Word2vec

Word2vec [88] is a 2-layer neural network whose goal is to represent tokens by the context in which they appear. We used the skip-gram architecture of word2vec, which receives single tokens as input and embeds them to predict nearby target tokens. The reason is that skip-gram works better with small datasets and has better representations of rare tokens [99]. Training samples were built from patient trajectory sequences by collecting all tokens that appear in a fixed-size context window and assigning them to the token in the center of the window. For our case, the context window extended over 5 tokens on both sides. During training, the model updates its parameters by maximizing the likelihood of predicting the context tokens given the corresponding input tokens. This is achieved by applying the softmax function to the dot product of the input token embedding and all possible token embeddings, which produces a probability distribution of the target context token over the vocabulary. After training, tokens that appear in similar contexts are expected to be close to each other in the embedding space, enabling word2vec to capture meaningful relationships between medical concepts.

#### 2.2.2 FastText

FastText [89, 90] is a neural network that extends the word2vec model by representing words as bags of character n-grams, capturing subword information. FastText can represent unseen words based on their subword units. The architecture and training schedule of fastText is similar to word2vec. The main difference is that, before being processed further, a token embedding is computed as the sum of the embeddings of its n-grams. The motivation to use fastText in our case is that codes from biomedical terminologies tend to be structured in a hierarchical manner (e.g., ICD10-CM, ICD-PCS, ATC). Subword information may hence automatically include this hierarchy in the token representations. To add subword information to the medical concept tokens, we used n-grams for 2 ≤ n ≤ 5. This means that any medical concept code is split into all possible n-grams with any n such that 2 ≤ n ≤ 5, plus the code itself. For example, the ICD10-CM code “S060X1” becomes {“S060X1”, “S0”, “06”, “60”, “0X”, “X1”, “S06”, “060”, “60X” “0X1”, “S060”, “060X”, “60X1”, “S060X”, “060X1”} before encoding.

#### 2.2.3 GloVe

GloVe [91] is a count-based algorithm that captures global word co-occurrence statistics. It constructs a co-occurrence matrix by counting how frequently words co-occur in the sequences of the dataset and then factorizes the matrix to obtain word embeddings. Unlike word2vec and fastText, GloVe considers the entire co-occurrence matrix during training, rather than focusing on local context windows around each word. In principle, this allows GloVe to capture more complex semantic and syntactic relationships between words that may be separated by several words or even sentences. In our experiments, to create the co-occurrence matrix, we counted how many times each pair of tokens appeared in a common patient trajectory sequence of the training dataset. Empirically, this approach led to better performance than using a fixed-size context window to compute co-occurrences. During training, the model embedded token pairs to predict their co-occurrence score.

#### 2.2.4 Creating training sequences

For fastText and word2vec, we followed the procedure of reference [83] to create the training sequences. The procedure shuffled the content of patient trajectory sequences when building center-context token pairs, which resulted in better predictive performance. The reason was that tokens prepended to the sequence, e.g., demographics, were otherwise almost never part of other tokens’ context windows. In the current work, to preserve local relationships among tokens in the learned representations, we shuffled each sequence only with 50% probability. We also ran an alternative training schedule, where no shuffling was applied, but all context windows artificially included the outcome tokens, demographic information, as well as the 3 most important diagnosis codes, in addition to the normal context tokens. However, this alternative training procedure produced very small and inconsistent improvements. For GloVe, as the co-occurrence matrix is created using the whole trajectory (i.e., window size equals to the trajectory length), we used the original trajectory sequence.

### 2.3 Visualization of medical concept embeddings

We visualized the embeddings of different medical concepts present in the MIMIC-IV dataset, as computed by the different language models described in section 2.2. More specifically, using the t-SNE algorithm [100], we generated a 2-dimensional representation of the embeddings of all ICD10-CM (diagnoses and reason for visits), ICD10-PCS (procedures) and ATC (medication) codes appearing in the model’s vocabulary. We used the python packages scikit-learn [101], Dask [102], and cuML [103] to implement the t-SNE algorithm and evaluate its outputs, with the following hyper-parameters: perplexity = 50.0, n_neighbours = 32, learning_rate = 200.0, metric = cosine, n_iter = 100,000.

To evaluate the capacity of each model’s embeddings to capture the semantic meaning of medical concepts, we labeled the codes with the main terminological subcategories to which they belong. Our expectations were that trained embedding would form clusters of medical concepts that correspond to these subcategories. For ICD10-CM codes, we used the ICD10 chapter indices^3^, which mainly correspond to the first letter of the code, and encode the general type of injury or disease, e.g., G (*nervous)*, I (*circulatory)*, or J (*respiratory)*. For ATC codes we used the first letter as well, which indicates the main anatomical group that the medication is intended to act upon, e.g., C (*cardiovascular system*), and D (*dermatologicals*). For ICD10-PCS codes, although the first character indicates the general category of the procedure, e.g., 7 (*osteopathic*) or B (*imaging*), most entries start with the character 0 (*medical and surgical procedures*). For this reason, in addition to first-letter subcategories, we split subcategory 0 using the second character, which stands for the body system or general anatomical region involved in the procedure, e.g., 0D (*medical and surgical procedures, gastrointestinal system*) or 0T (*medical and surgical procedures, urinary system*). Note that we only kept subcategories that, on the one hand, held at least 1% of the codes in the terminology and, on the other hand, represented at least 1% of the tokens of the training dataset. More details about all subcategories are available in the supplementary information, Appendix A2.

### 2.4 Prediction tasks

We extrinsically evaluated the quality of embeddings produced by the trained models (see section 2.2) with two types of patient trajectory prediction tasks (Figure 3, left): binary and multi-label. For the binary task, the aim was to predict the outcome of a stay, namely length-of-stay (normal or extended), readmission, and mortality. For the multi-label task, the aim was to predict the clinical events in the patient trajectory for a given category, namely diagnoses or reasons for visit (ICD10-CM), procedures (ICD10-PCS), and medications (ATC). Importantly, no model supervision was involved, i.e., predictions were solely based on the cosine-similarity between embeddings of patient trajectories and possible target token (Figure 3, right). More specifically, for each task, we assigned a score to any code belonging to the relevant category, i.e., the outcome labels for the binary prediction task, and all codes of the specific category (ICD10-CM, ICD10-PCS, ATC) for the multi-label prediction task. The score was defined as 1.0 minus the cosine-similarity, which we used to compute the receiver operating characteristic and the precision-recall curves for any combination of task and model. To quantify the alignment between patient trajectories and target medical codes with more granularity, model inputs included only a fraction of the patient trajectory sequences. Starting from demographics tokens only, we gradually increased the proportion of medical tokens given to the model (i.e., P = 0.0, 0.1, 0.3, 0.6, 1.0). We give more details about each prediction task in sections 2.4.1 and 2.4.2.

**Figure 3.**
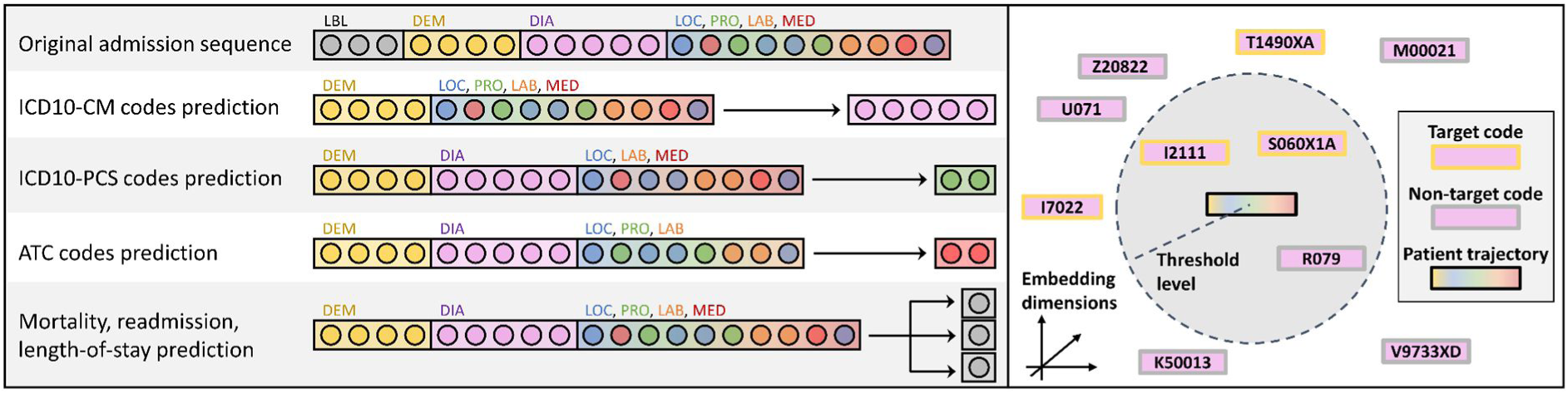
Prediction tasks performed by the models. Left. Patient trajectory sequences from the testing dataset were embedded by the trained language models. Task-specific target tokens were removed from the input sequences embedded by the models. Label tokens (used for the binary prediction tasks) were also removed for any prediction task. Right. For each predicted category, patient trajectory embeddings were compared to embeddings of any code belonging to that category (ICD10-CM codes in the figure example). Model predictions were computed as the set of medical concept tokens that are within a threshold level of dissimilarity from the patient trajectory embedding (note that, for illustration purpose, the figure depicts a threshold Euclidean distance, but we actually used 1.0 minus cosine-similarity). Predicted target tokens within threshold level correspond to true positives and predicted non-target tokens to false positives. Target tokens outside threshold level correspond to false negatives. Based on these numbers, precision-recall and ROC curves were computed using different thresholds.

#### 2.4.1 Length-of-stay, readmission, and mortality prediction

In these tasks, single label tokens for length-of-stay, readmission, and mortality were predicted by the model, based on patient trajectory embeddings. Each label token had two possible statuses: *normal* and *extended* for length-of-stay; *no-readmission* and *readmitted* for readmission; *alive* and *dead* for mortality. During training, these tokens were added at the beginning of the patient trajectory sequences and, hence, models learned to represent these tokens in relation to other medical concepts. During evaluation, these tokens were removed from the trajectory sequence (Figure 3, left). The prediction of the model was then simply computed as the outcome token whose embedding is closer to the patient trajectory embedding in terms of cosine-similarity (Figure 3, right). For each task, we evaluated model prediction performance with the areas under the receiver operating characteristic (AUROC) and the precision-recall (AUPRC) curves.

#### 2.4.2 Prediction of diagnoses and reasons for visits, procedures, and medications

In these tasks, diagnosis, and reason for visit (ICD10-CM), procedure (ICD10-PCS) and medication (ATC) codes were predicted by the model based on patient trajectory embeddings. For each task, the tokens belonging to the category of tokens being predicted were removed from the input sentence and set as target tokens (see Figure 3, left). Outcome tokens, i.e., length-of-stay, readmission, and mortality, were also removed from the input. Cosine-similarity was then computed between the embedding of the input sequence and any potential target token. These tasks are particularly challenging. For example, when predicting ICD10-CM codes, the model must correctly guess, on average, 11 tokens, any of which comes from a set of several thousand possible values. We evaluated model prediction performance for different levels of lenience. First, we considered a model prediction as a hit by requiring an exact match between predicted and target tokens. Then, we only asked for a lenient match, i.e., having N letters in common with at least one of the target tokens, for N = 1, 2, 3, and 4. Finally, precision, recall, and AUPRC were computed for each level of lenience.

## 3. Results

### 3.1 Qualitative visualization of medical concept embeddings

This section provides insights into how different models capture semantic and hierarchical structures within medical terminologies. To this end, we visualized ICD10-CM, ICD10-PCS, and ATC code embeddings as produced by word2vec, fastText, and GloVe, after reducing their dimensionality using the t-SNE algorithm. To quantify embeddings’ ability to represent medical concepts, we used the following metrics.

– Rate reduction [104, 105] measures the ability of a representation to compress information by calculating the difference between the rate distortion of the entire dataset and the mean rate distortion of each class when considered separately. Rate distortion quantifies the number of bits needed to encode any representation. A higher rate reduction indicates a more efficient representation that distinguishes between different classes with less information loss.
– Homogeneity evaluates the output of a clustering algorithm, as the extent to which each cluster contains only members of a single class. It reflects how well clusters derived from model embeddings align with the true categories in the data.
– Completeness assesses whether all members of a given class are assigned to the same cluster. It reflects how well clusters derived from model embeddings capture the cohesion of each class within the embedding space.
– The V-measure is the harmonic mean of homogeneity and completeness. It serves as an overall indicator of the quality of the clusters derived from model embeddings.

#### 3.1.1 Comparison to ICD10-CM, ICD10-PCS, ATC terminologies

Figure 5 shows the output of the t-SNE algorithm using embeddings obtained by the word2vec, fastText, and GloVe models for the ICD10-CM, ICD10-PCS, and ATC codes. Note that for ICD10-CM codes we did not visualize the subcategory R, since it stands for “symptoms, signs and abnormal clinical and laboratory findings, not elsewhere classified”, which we deemed too heterogeneous to have any chance to form a cluster. Moreover, we separated the subcategory “injury, poisoning and certain other consequences of external causes” into two, i.e., S and T (instead of keeping them together).

**Figure 4.**
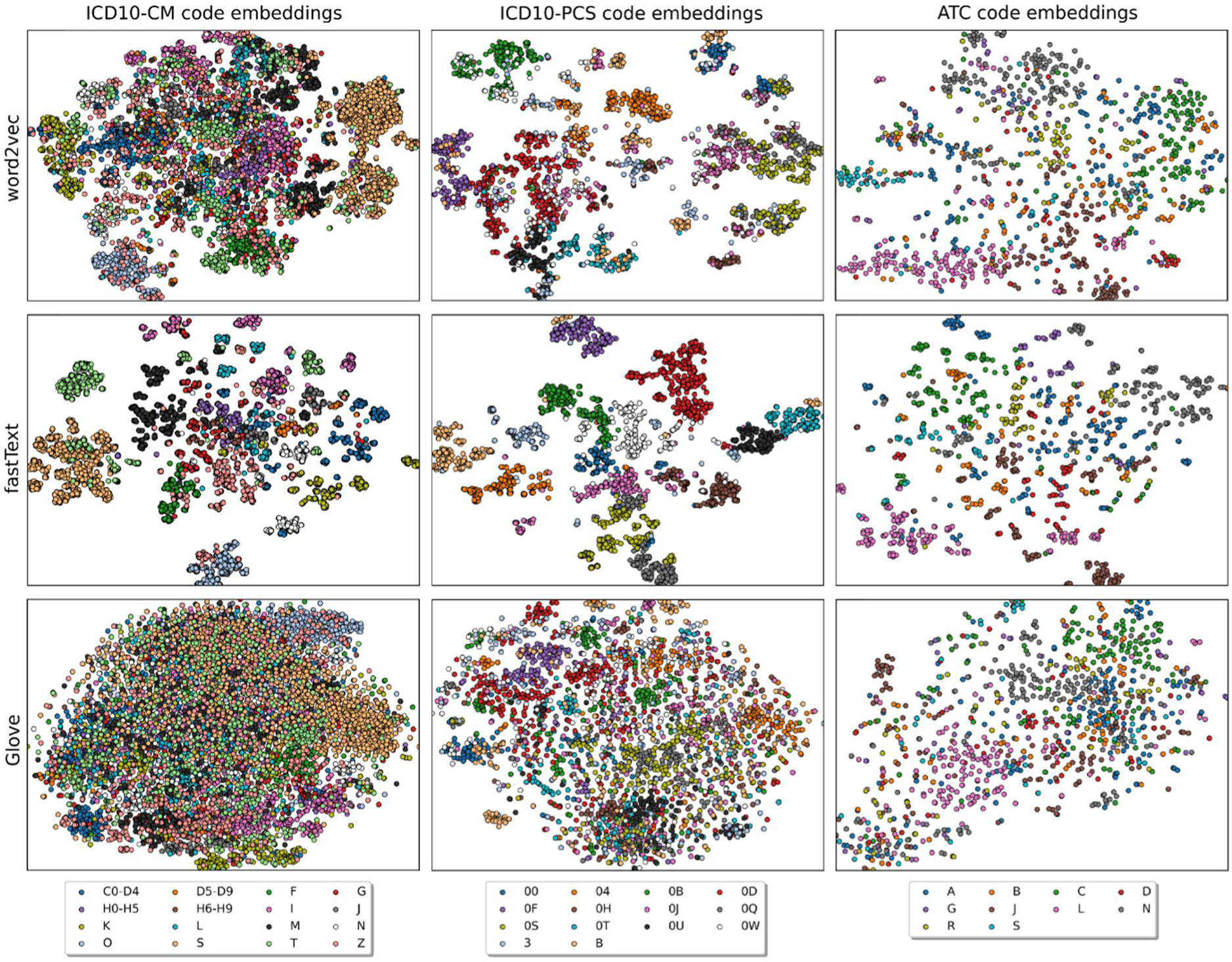
Medical concept embeddings obtained after training the language models. We used the t-SNE algorithm to reduce the dimensionality of embeddings (d = 2). Colors represent the main subcategories of ICD10-CM, ICD10-PCS, and ATC codes (for details, see section 2.3 and supplementary information, Appendix A2).

**Figure 5.**
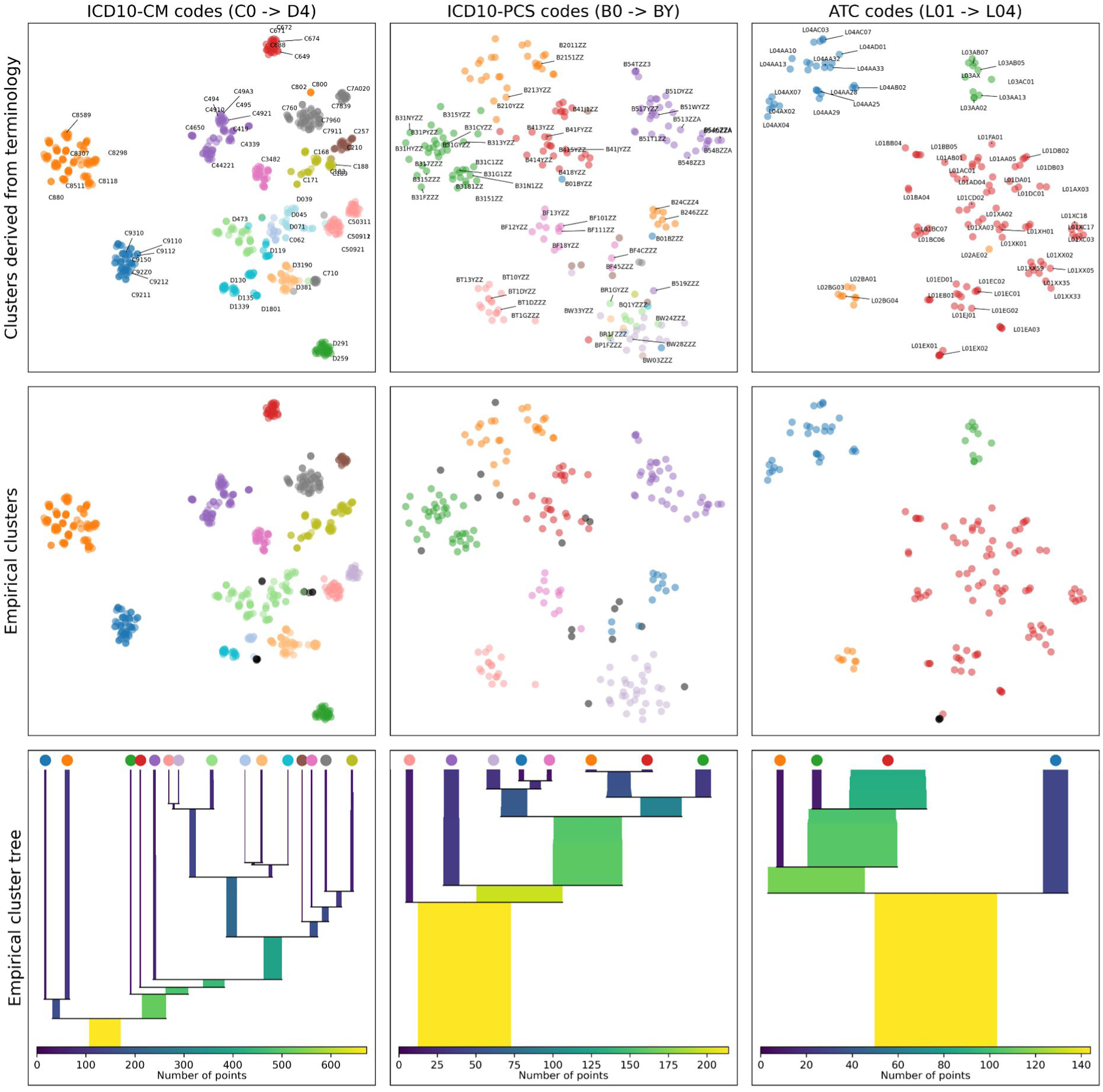
Clusterization of a selected subset of medical concepts using fastText embeddings after dimensionality reduction (t-SNE, d = 2). Top. Theoretical clusters. Samples are positioned following reduced embeddings. Colors are derived from the second hierarchical level of ICD10-CM, ICD10-PCS, and ATC codes. Center. Empirical clusters. Colors are assigned using the hdbscan clustering algorithm, and then aligned with the top row by maximizing the number of sample matches between theoretical and empirical clusters. Samples labeled with a black color were not assigned to any cluster (i.e., labeled as noise). Bottom. The empirical cluster trees were also generated with hdbscan. We cut the tree after the number of theoretical clusters was reached (i.e., 15 for ICD10-CM, 8 for ICD10-PCS and 4 for ATC codes). We added color patches that correspond to the empirical cluster in the center row.

Amongst all models, fastText provides embeddings whose t-SNE visualization forms the most separate clusters. Moreover, embedding clusters are visually aligned with the subcategories that are defined in section 2.1, denoted by the different colors in Figure 4. Since the t-SNE algorithm preserves local distance relationships between data samples, this suggests that semantic relationships between medical codes are part of the low-level structure of fastText embeddings. For ICD10-CM codes, the subcategories whose fastText embeddings are the most spread out and undefined are G (*diseases of the nervous system*), L (*diseases of the skin and subcutaneous tissue*), and H6-H9 (*diseases of the ear and mastoid process*). Some codes have a substructure within subcategories, e.g., N (*diseases of the genitourinary system*) which presents two subclusters that, upon further inspection, separate conditions related to male and female genital organs. Other subcategories are mixed but are still part of well-defined clusters. For example, ICD10-CM tokens of subcategories S and T (*injury, poisoning and certain other consequences of external causes*) are separated into one cluster made only of T-tokens, and another one that mixes S and T tokens. Upon closer examination, the cluster composed of both subcategories corresponds to injuries made to an external body part (i.e., fractures, burnings, frostbites, etc., included in ICD10-CM codes that start with S00 to T34 and T66 to T88), while the other cluster corresponds to internal causes of harm (toxic substance effects, poisoning, etc., included in ICD10-CM codes that start with T36 to T65). This suggests that, even though subword information (in this case, the first letter of ICD10-CM codes) acts as prior knowledge during training in the case of fastText, data-driven statistics are smoothly integrated into the medical embedding representation of the trained model. In general, fastText embeddings of all subcategories tend to have an inner substructure. To compare these substructures to the second hierarchical level of terminologies, we also performed a clustering analysis of fastText embeddings, focusing on a specific set of subcategories (see section 3.1.2). Word2vec produces embeddings that are significantly more mixed between subcategories, especially for ICD10-CM codes. For example, subcategory T, which forms well defined clusters with fastText, spreads across the entire representation space. For ICD10-PCS codes, embeddings are more aligned with subcategories, but not as much as using fastText. For example, subcategories 0H, 0J, 0Q, 0S (standing for *medical and surgical procedures performed on the skin, subcutaneous tissues, bones, and joints*, respectively), are found to be mixed with each other, whereas they form distinct clusters with fastText. Another example is the subcategory 0W (*Medical and Surgical Body Systems – Anatomical Regions, General*), for which tokens are completely shattered when provided by word2vec, which is not the case for fastText. Finally, GloVe embeddings are the ones that are the most mixed between subcategories after t-SNE reduction.

To quantify the quality of the embeddings learned by the models, we computed rate reduction [104, 105] for any combination of model representation and medical concept category. Table 2 shows rate reduction using the full dimensionality of the embeddings or the output of the t-SNE algorithm (i.e., a 2-dimensional representation).

**Table 2.** Rate reduction of medical concept representations. Higher values mean better representations (note that rate reduction cannot be compared across dimensionalities, since it depends on the dimensionality). When using reduced embeddings (dim = 2), fastText obtains higher rate reductions. This reflects its better alignment with the first level of biomedical terminologies. When using raw model embeddings, GloVe tends to obtain slightly larger rate reductions.

|  |  | word2vec | fastText | GloVe |
| --- | --- | --- | --- | --- |
| Raw embeddings<br>(d = 512) | ICD10-CM | 410 | **531** | 403 |
|  | ICD10-PCS | 1387 | 1245 | **1463** |
|  | ATC | 1804 | 1504 | **1833** |
| Reduced embeddings<br>(t-SNE, d = 2) | ICD10-CM | 0.38 | **0.72** | 0.11 |
|  | ICD10-PCS | 0.47 | **1.20** | 0.09 |
|  | ATC | 0.21 | **0.30** | 0.07 |

When considering embeddings after dimensionality reduction, fastText obtains the largest rate reduction score for all categories (Table 2, d = 2), which reflects the better alignments of reduced fastText embeddings with the hierarchy of biomedical terminologies (Figure 4). For unreduced embeddings, while fastText achieves the largest rate reduction score for ICD10-CM codes, GloVe shows slightly larger scores for ICD10-PCS and ATC codes (Table 2, d = 512). This suggests that, if semantic information of medical concepts is accurately represented in the embedding space of GloVe, they are part of a more global and high-dimensional structure. This may have an impact on the quality of embeddings for prediction tasks (see section 3.2).

#### 3.1.2 Hierarchical analysis

We assessed the ability of all models to uncover the hierarchical relationships among ICD10-CM, ICD10-PCS, and ATC codes. Specifically, we evaluated whether models’ embeddings could be used to rediscover the second hierarchical level of medical terminologies. We focused on ICD10-CM codes beginning with C0 to D4, ICD10-PCS codes beginning with B0 to BY, and ATC codes beginning with L01 to L04. We generated empirical clusters from the reduced (2-dimensional) embedding vectors of these codes and compared them to theoretical clusters based on the second hierarchical level of the terminologies, namely the second character of ICD10-CM and ICD10-PCS codes, and the third character of ATC codes. We used the hdbscan algorithm [106] to compute the empirical clusters, either using raw embeddings or embeddings after t-SNE dimensionality reduction. Note that we used the number of theoretical clusters as prior knowledge when determining the clusters (i.e., we set a flat lambda value cutoff in the hdbscan cluster tree to obtain the same number of clusters as in the medical terminology). We measured the quality of the clusters generated by each model (Table 3).

**Table 3.**
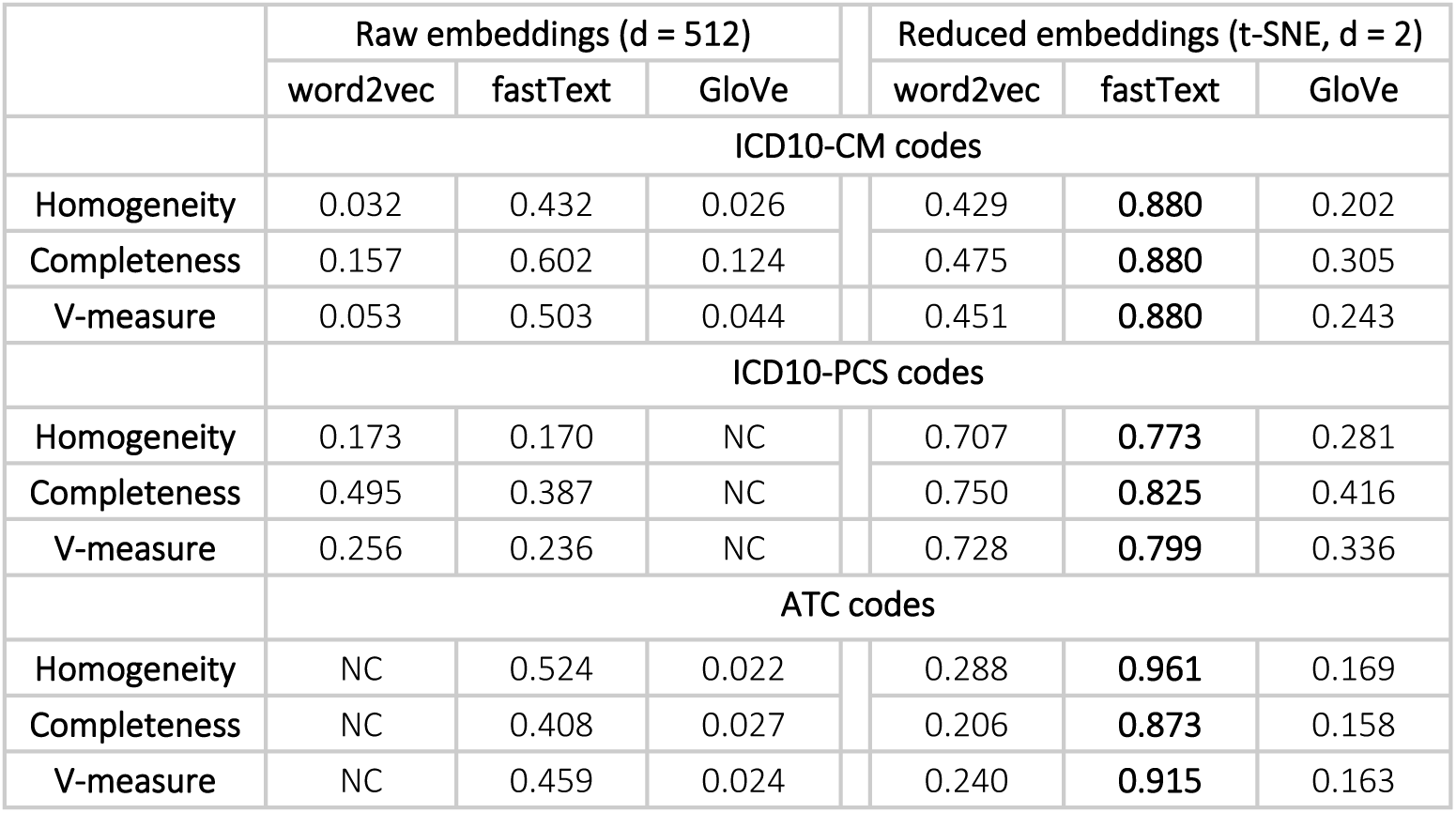
Evaluation of empirical clusters as determined by model embeddings. We quantified the homogeneity of empirical clusters and their match with theoretical clusters (completeness). V-measure is computed as the harmonic mean between homogeneity and completeness. NC: the clustering algorithm could not identify any cluster.

Reduced fastText embeddings consistently outperform other combinations when used to rediscover the second level of the hierarchy of biomedical terminologies. For this reason, we visualized theoretical clusters, as well as empirical clusters generated with this combination of model and dimensionality (Figure 5).

The empirical clusters derived from fastText embeddings are generally well-aligned with the theoretical clusters. Still, empirical data proposes alternative ways of defining the terminology of medical codes by identifying new clusters. For example, procedure codes at the bottom-right of the diagram (Figure 5, center column, top vs. center row) form a well-defined group although the medical concepts belonging to that cluster come mainly from two different ICD10-PCS subgroups, namely BW (*anatomical regions*) and BR (*axial skeleton, except skull and facial bones*). Moreover, some singular medical concepts are moved to different clusters by empirical data. For example, for ATC codes, L02AE02 (*leuprorelin*) that belongs to the orange theoretical cluster (L02: *endocrine therapy*) is moved towards the blue empirical cluster (L01: *antineoplastic agents*) by empirical data, which is also a valid categorization of *leuprorelin*, i.e., an antineoplastic agent (Figure 5, top-right).

Also note that, for ICD10-CM codes, the way we defined theoretical groups (i.e., relying on the second code letter) includes inaccuracies. For example, the group *Malignant neoplasms of ill-defined, other secondary and unspecified sites* consists of ICD10-CM codes that start with C76 to C80, which include samples coming from both theoretical clusters C7 (gray) and C8 (orange). Empirical clusters correctly identified this issue (Figure 5, top-left, group of orange codes around the gray cluster), even though subword information should bias C80 tokens to be closer to other tokens starting with C8. Another example for ICD10-CM codes are the groups *malignant neoplasms of breast* (codes that start with C50) and *malignant neoplasms of female genital organs* (code that start with C51 to C58). Empirical embeddings correctly identified these two groups (Figure 5, center row, right column, mauve and salmon clusters on the right of the plot), even though they belong to the same theoretical cluster C5.

### 3.2 Prediction tasks

In this section, we analyze the application of word2vec, fastText, and GloVe for various prediction tasks using medical concept embeddings. These tasks include predicting length-of-stay, readmission, and mortality outcomes, as well as diagnosis, procedures, and medication codes from patient trajectory embeddings. Model predictions are based on the proximity of medical concepts to patient trajectory embeddings, i.e., generated in an unsupervised manner. To evaluate model performance, the following metrics are used.

#### 3.2.1 Length-of-stay, readmission, and mortality prediction

Figure 6 shows AUROC and AUPRC obtained with the embeddings of any model, when predicting outcomes from patient trajectories. Note that AUPRC is a better measure for prediction problems with unbalanced classes, which is our case. Also note that P is the fraction of the tokens of each patient trajectory sequence given to the model as an input for prediction, increasing from P = 0.0 (demographics tokens only) to P = 1.0 (full trajectory). First, increasing the fraction of tokens given as input to the models always improves their prediction performance, for any model. However, fastText seems to benefit slightly less from more information about patient trajectories, as its performance peaks to lower values than what is obtained with word2vec and GloVe, even though they all start from similar baseline performances at P = 0.0.

**Figure 6.**
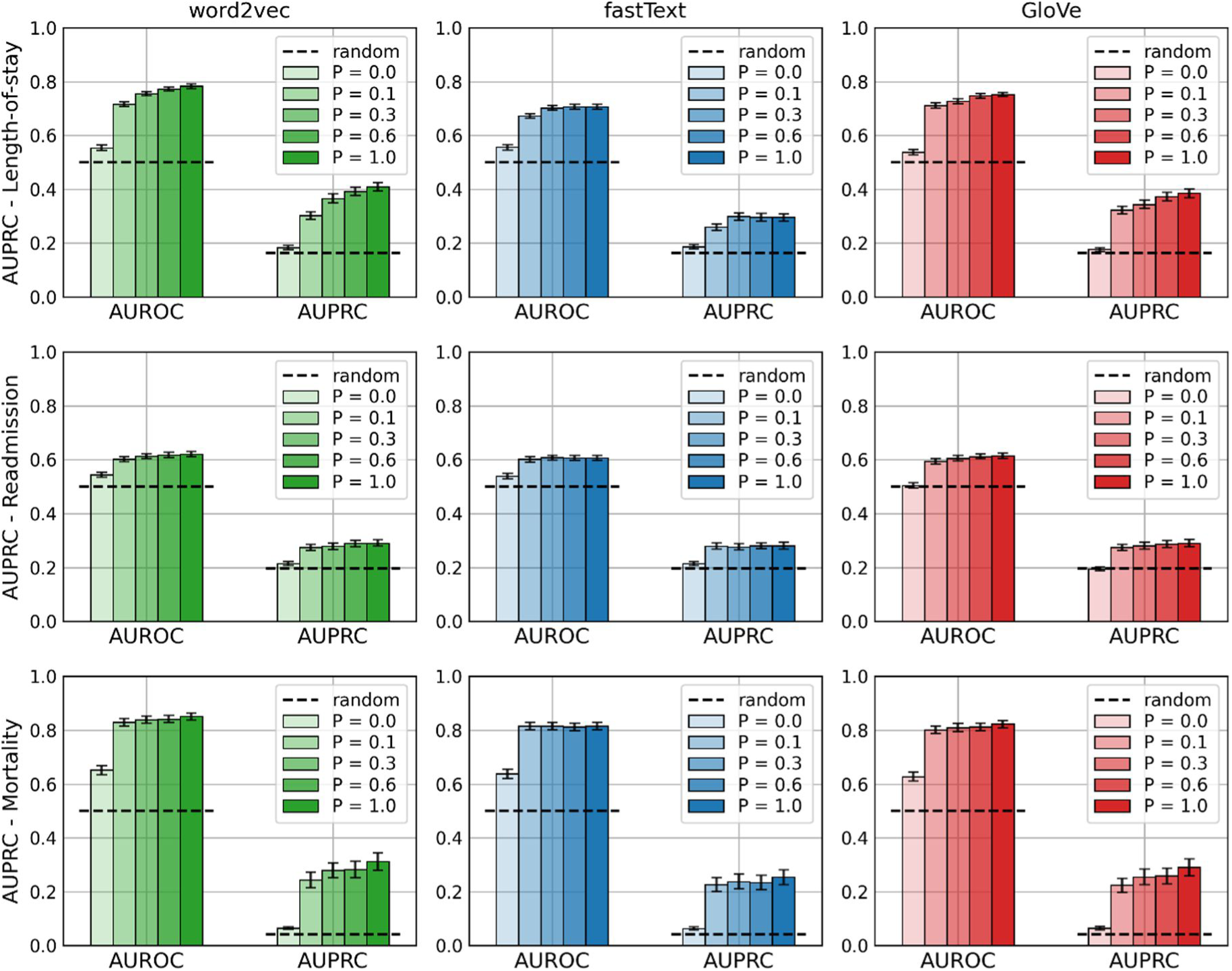
AUROC and AUPRC obtained with word2vec, fastText and GloVe for outcome prediction tasks. P stands for the different proportions of the patient stay tokens given as input to the models (see section 2.4). The random performance for AUPRC corresponds to the number of positive samples over the total number of samples. The reported means and standard deviations were calculated using the Bag of Little Bootstraps algorithm [107, 108] with n_bootstraps = 100, n_subsamples = 100, subsample_size = 10,000.

#### 3.2.2 Comparison of patient outcome embeddings

In Figure 7, we visualize patient trajectory embeddings for the different outcomes – length-of-stay, readmission, and mortality – and compare them with the outcome token embeddings themselves to assess whether high-level outcome information is present in the local structure of the embeddings. Embeddings for each patient trajectory in the testing dataset were generated by removing the outcome labels and averaging the remaining concept embeddings. The 2-dimensional representations were then obtained by applying the t-SNE algorithm for word2vec, fastText, and GloVe embeddings to the set of patient trajectory embeddings to which we added all medical concept embeddings from the vocabulary. For a better comparison between medical concepts and patient trajectories, we transformed all embeddings prior to t-SNE dimensionality reduction using a representational dissimilarity matrix (RDM) [109]. RDM represents each embedding by its pattern of dissimilarities to all other embeddings in the dataset. To compute dissimilarity, we used the correlation distance. Results show that antagonistic outcome tokens tend to reach different locations, i.e., have different patterns of dissimilarities with patient trajectories. One exception is the case of antagonistic readmission outcome tokens, whose locations are much more similar, for all models. This aligns with the finding that the readmission prediction task achieves lower levels of performance compared to random predictions (Figure 6, second row). Moreover, positive patients in the mortality prediction case seem to be confined to a more specific region than for other outcome categories and disposed around the corresponding outcome token. This aligns with the finding that models achieve slightly higher mortality prediction performance compared to random predictions (Figure 6, last row).

**Figure 7.**
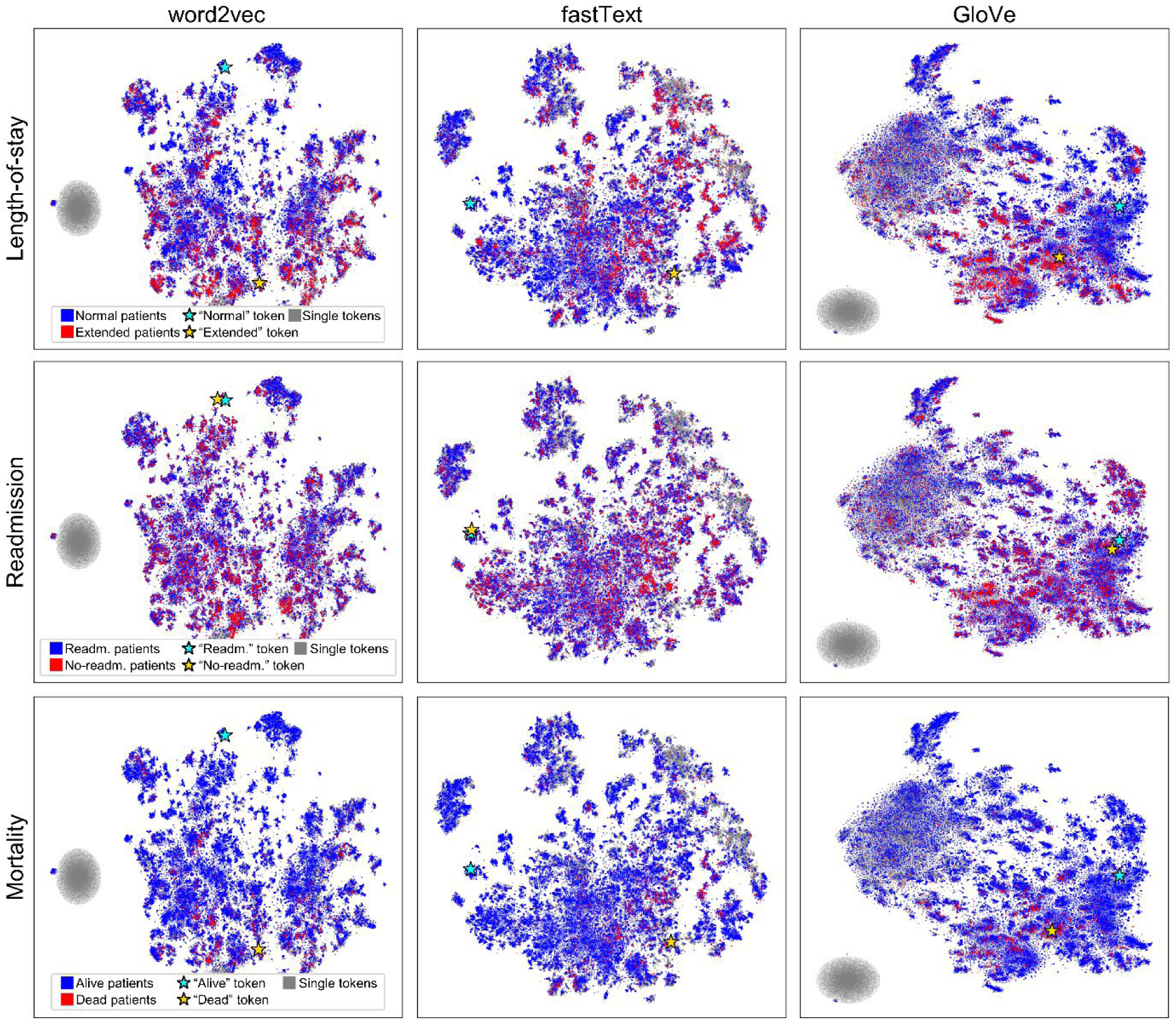
Visualization of patient trajectory embeddings for all combinations of different outcomes (length-of-stay, readmission, mortality) and models (word2vec, fastText, GloVe), after t-SNE dimensionality reduction. Patient trajectory embeddings are labelled by outcome (blue vs. red). We also added the reduced embedding of all medical codes in grey and highlighted the specific outcome tokens using star symbols. Note that the grey blobs in the word2vec and GloVe visualizations correspond to the medical codes from the testing dataset which are not part of the models’ vocabularies and assigned the same “unknown” encoding (reaching slightly different locations because of the stochasticity of the t-SNE algorithm). This does not happen for fastText, which uses n-gram information to represent unknown tokens.

#### 3.2.3 Diagnosis, procedure, and medication code prediction

Figure 8 shows the AUPRC obtained with the models when predicting all medical codes of a category given the patient trajectory embeddings (AUROC values are shown in the supplementary information, Appendix A3). First, word2vec has, in most cases, a lower score than fastText. Second, GloVe embeddings tend to outperform other models when P = 0.0 (i.e., using only demographic tokens), whereas fastText tends to outperform other models as the level of partial information increases. We include two supplementary figures in which we directly compare model performance for different levels of partial information (Appendix A4 for AUPRC, Appendix A5 for AUROC). The results show that, for word2vec and fastText, model performance improves as more information (higher P) is available, whereas performance worsens for GloVe. Overall (i.e., across all levels of P), GloVe obtains the best performance for the diagnosis and medication code prediction tasks (e.g., 0.45 and 0.81 AUPRC for 1-letter match lenience), whereas fastText is the best model for the procedure code prediction task (e.g., 0.66 AUPRC for 1-letter match lenience).

**Figure 8.**
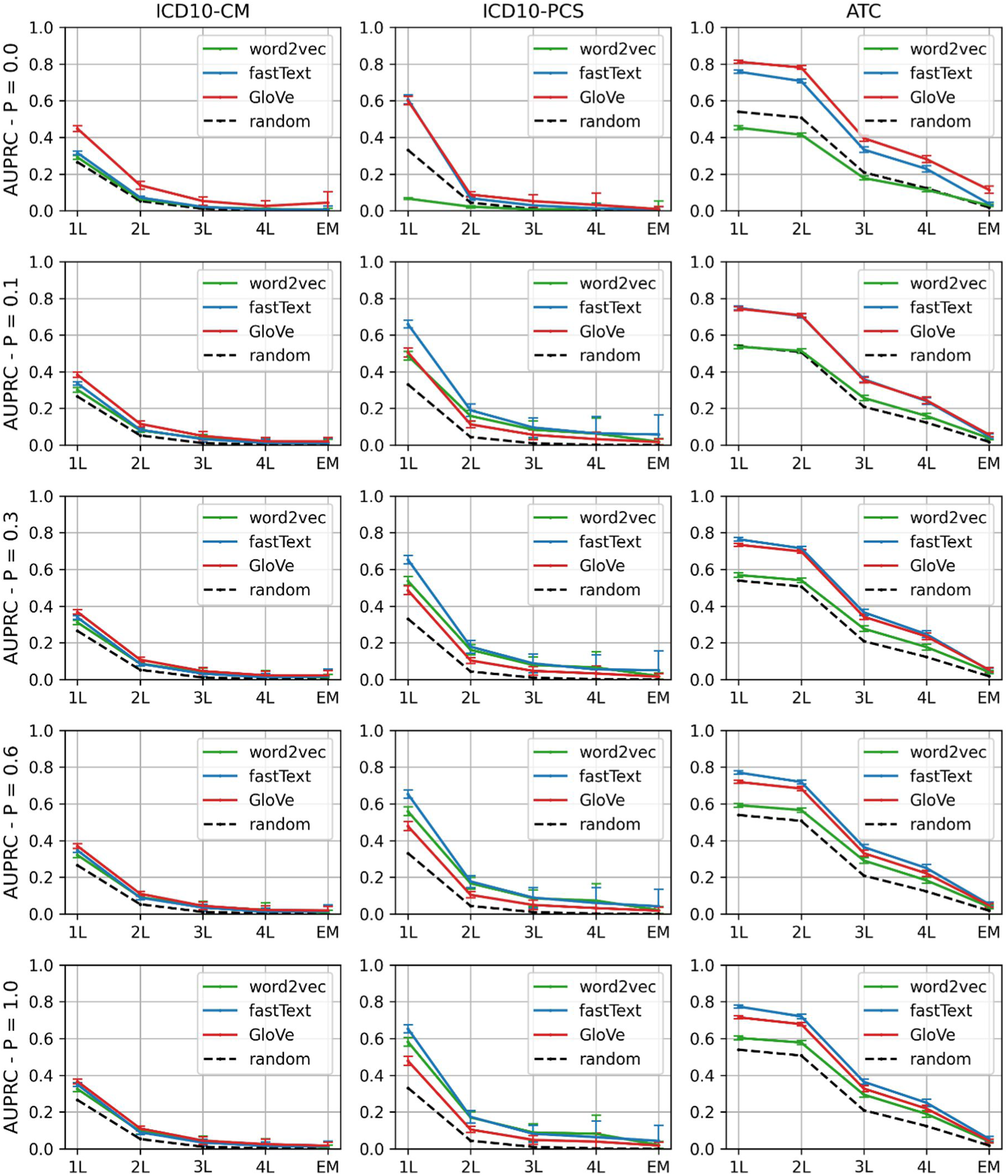
AUPRC obtained with word2vec, fastText and GloVe for medical concept prediction tasks. L stands for the lenient letter match. For example, 1L means that precision and recall were computed on the basis of the first letter of the codes only. EM means that an exact match was required between a model prediction and a target code. P stands for the different proportions of the patient trajectories given as input to the models (see section 2.4). The random performance corresponds to the number of positive samples over the total number of samples. The reported means and standard deviations were calculated using the Bag of Little Bootstraps algorithm [107, 108] with n_bootstraps = 100, n_subsamples = 100, subsample_size = 10,000.

We also performed medical code prediction in a more realistic setting. For ICD10-CM codes, patient trajectory embeddings were used to predict only the most important diagnosis code (see Figure 1, p1), given the full patient trajectory from which all outcome and diagnosis tokens were removed. For ICD10-PCS and ATC codes, we used the embeddings of a patient’s trajectory, including events only up to a certain point in time, to predict the next occurring procedure or medication token. Figure 9 shows AUPRC for these prediction tasks (AUROC values are shown in the supplementary information, Appendix A6). These tasks are still challenging for the models (performance levels are not higher than in Figure 8). We assume that this is due to the lack of temporal information encoded by the embeddings, that is, they tend to learn hierarchical relationships among medical concepts but ignore their sequential relationships, which is trajectory dependent. Overall, word2vec reaches the best performance for the most important diagnosis code prediction, whereas fastText is the best model for the next procedure and medication code prediction task.

**Figure 9.**
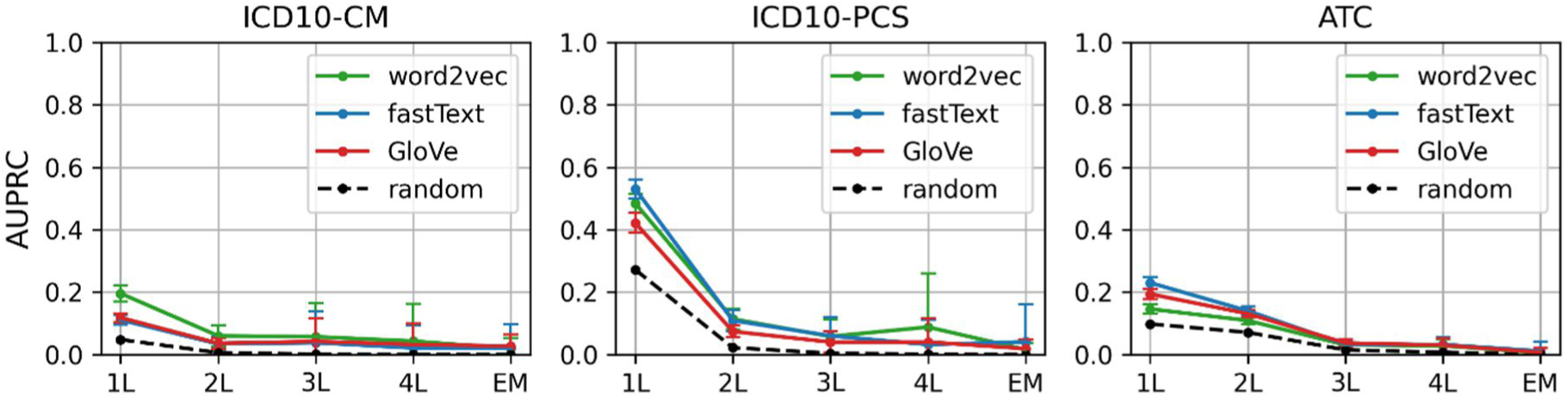
AUPRC obtained with word2vec, fastText and GloVe for alternative prediction tasks. We used model embeddings to predict the most important ICD10-CM code of the patient trajectory, as well as the next ICD10-PCS and ATC code, given the medical events that happened up to the target code. L stands for the lenient letter match. For example, 1L means that precision and recall were computed on the basis of the first letter of the codes only. EM means that an exact match was required between a model prediction and a target code. The reported means and standard deviations were calculated using the Bag of Little Bootstraps algorithm [107, 108] with n_bootstraps = 100, n_subsamples = 100, subsample_size = 10,000.

## 4. Discussion

In this study, we compared the ability of different language models, word2vec, fastText, and GloVe, to represent medical concepts by expressing patient trajectories as sequences of medical codes. Using dimensionality reduction and clustering algorithms, we showed that language models can learn data-driven embeddings that retrieve the semantic meaning of medical concepts, as provided by biomedical terminologies. These detailed visualizations and interpretative analyses serve as a valuable resource for researchers interested in deeper explorations of the interpretability of language models in the medical field. Using an unsupervised approach, we also compared the respective capabilities and limitations of different language models for patient trajectory prediction.

To evaluate the semantic content of embeddings produced by the different language models, we explicitly compared their alignment with existing medical concept terminologies. Using rate reduction as a quantitative measurement, we showed that, in terms of low-level representation, fastText is the model whose embeddings are best aligned with the hierarchy of existing medical concept terminologies (Figure 4 and Table 2). Note that, in the case of fastText, subword information includes the hierarchical structure of medical terminologies. Indeed, some subwords of ICD10 and ATC codes correspond to their top-level categories. For instance, the subwords of the ICD10-CM code T3302XA (*superficial frostbite of nose, initial encounter*) include T33 (*superficial frostbite*) and T3302 (*superficial frostbite of nose*). However, most subword tokens are irrelevant, such as 02XA or 330 in the aforementioned example. Moreover, the hierarchy of medical terminologies does not strictly follow subword information. For example, medical concepts describing physical injuries consist of ICD10-CM codes that start with S00 to T34, and from T66 to T88, skipping codes that go from T35 to T65. Still, fastText embeddings are able to update prior subword knowledge and uncover the complex hierarchy of medical terminologies (Figure 4 and 5). This suggests that fastText is an efficient way of combining prior knowledge (included in subword information, which reflects existing terminologies) with data-driven representations (which depend on the history of patients at the ICUs).

We extrinsically evaluated the quality of the medical concept embeddings via outcome and trajectory prediction tasks using an unsupervised method. First, we performed a multi-label prediction task in which tokens belonging to one category (ICD10-CM, ICD10-PCS, or ATC codes) were detached from patient trajectories and used as target tokens. Patient trajectories were embedded using the language models (Figure 2) and compared to the embeddings of the target tokens using cosine-similarity (Figure 3). In terms of multi-label prediction (Figure 8), GloVe tends to outperform fastText and word2vec for low levels of partial information (especially when only demographic tokens are available, i.e., P = 0.0) but worsens as P increases. We interpret this as the result of GloVe long-range co-occurrence statistics being able to extract useful patient features from even limited data, but more sensitive to averaging many medical tokens, which can add noise to these statistics. When more data is available, word2vec and fastText performances increase (see supplementary information, Appendix A4 and Appendix A5), meaning that adding more medical concept codes improve their representation of patient trajectories. We suggest this is due to the good alignment between these models’ medical concept embeddings and medical terminologies (see Figure 4), which ensures that added tokens contribute to relevant and specific clinical information and improve the accuracy of the patient trajectory representation. In line with this interpretation, fastText obtains the best performance for the next procedure and medication code prediction tasks (Figure 9). Predicting the next medical event may arguably require more fine-grained features about patient trajectories, and embeddings that align well with medical terminologies might be more prone to provide such features.

We also carried out a binary prediction task in which patient trajectory embeddings were compared to the embeddings of binary clinical outcome tokens (length-of-stay, readmission, mortality) that were prepended to each patient trajectory during training. As for the multi-label prediction tasks, we used an unsupervised method to generate predictions, which were computed as the outcome tokens that were more similar to the patient trajectory embeddings. First and foremost, achieved performance levels for any model are considerably low, with most AUPRC measurements rarely exceeding a value of 0.4. This was to expect given the simplicity of our unsupervised prediction method, as compared to the target outcomes which inherently depend both on global trajectory patterns and on nuanced interactions between medical concepts. Moreover, pooling all tokens together when building patient trajectory embeddings is likely suboptimal for capturing the rich, dynamic information contained in patient trajectories. A supervised method would probably be more suited to perform outcome predictions. This is reflected in Figure 7, where we visualized patient trajectory embeddings for different outcomes. These should ideally span contrasting regions of the embedding space, but actually present a large degree of overlap (at least in the reduced embeddings space). One exception is the mortality prediction task, in which positive patient trajectories are more confined and closer to the corresponding outcome token, which correlates with the slightly higher model performance for this category (see Figure 6, last row). Besides, all positive outcome tokens (i.e., tokens for readmission, extended stay, and death) tend to be very similar to each other, which might be a byproduct of prepending them to each patient trajectory during training, since they always appear in similar contexts.

Nevertheless, the binary outcome prediction task was used as a comparative evaluation method of model embeddings and their potential for more complex and high-level tasks. From our analysis, we found that word2vec and GloVe have similar performance levels, while they tend to outperform fastText (Figure 6). We suggest that the cause of this discrepancy lies in fastText’s intrinsic modeling of the hierarchical structure of medical terminologies. While this feature benefits fastText in the semantic alignment of its embeddings with existing medical terminologies, it seems to hinder the model in more high-level tasks, such as clinical outcome prediction. This task requires a broad and flexible representation of medical concepts and may be disrupted by fastText’s stricter adherence to the hierarchy of biomedical terminologies. This underlines the importance of understanding the specific strengths and weaknesses of each model in relation to the nature of the task at hand.

This study has several limitations. First, we focused only on methods that produce static embeddings. However, more modern language models, such as those based on attention, provide contextualized embeddings, which usually improve performance in downstream tasks [110, 111]. In this case, instead of having a unique representation, clinical concepts have many according to the context in which they appear. Contextualized representation would thus require a different clustering and concept mapping methodology and should be a subject of further study. Second, for all prediction tasks that we used to evaluate model embeddings, no supervision was involved. While supervised learning tends to improve upon unsupervised methods, our goal here was to compare the quality of the extracted embeddings rather than devise an optimal trajectory prediction method. In that sense, we argue that the unsupervised methodology provides a less biased comparison, as it is independent of the learning model. A supervised methodology based on the proposed embedding strategy could also be a topic for further research and would benefit from the results presented in the current study. Third, due to the lack of explicitly time-stamped information for diagnosis codes, i.e., clinical evaluation time instead of billing time, we used the diagnosis priority to order these codes in the patient trajectory. As patients tend to have a recoverable condition before ICU admission, these codes were inserted early during the original patient trajectory sequence generation. However, this may not necessarily always reflect the actual trajectory, as new diagnosis codes can be assigned to a patient during the ICU stay [112]. Fourth, there may exist 1-to-1 relationships between medical concepts, e.g., an antiretroviral therapy might be specific to HIV/AIDS patients. Hence, for the ICD10-CM, ICD10-PCS and ATC code prediction tasks, there might be some leakage from input to target tokens. Nevertheless, given the limited performance of the predictions, we assume that this effect is negligible. Finally, our study has a similar methodology to our previous work [83] in training language models with patient trajectories. Still, our current work stands out by its comprehensive comparison of multiple neural language models, novel evaluation methods using clustering algorithms for alignment with biomedical terminologies and analysis of both patient outcome and patient trajectory prediction, with new insights into medical data semantics and machine learning applications in healthcare. Note that Phe2vec [69] also shares a similar methodology, but its primary goal, building patient cohorts around specific diseases, diverges from ours.

## 5. Conclusion

We assessed the capabilities of different language models (word2vec, fastText, and GloVe), each coming with their own set of hypotheses, in expressing patient trajectories as sequences of medical codes. We found that these models can indeed learn data-driven embeddings that capture the semantic meaning of medical concepts. However, the effectiveness of these models varies based on the task at hand. While fastText aligns well with existing medical terminologies thanks to subword information, GloVe is more useful for more high-level tasks, such as clinical outcome prediction, thanks to its ability to consider long-range relationships and global co-occurrences in patient trajectories. These results offer important insights for supervised medical concepts and clinical outcomes prediction methods and open up several exciting avenues for future exploration. One promising research avenue is refining strategies for encoding subword information for representing medical concepts. For instance, a tokenization that aligns with ICD10 and ATC hierarchies, instead of relying on basic n-grams, could enhance the accuracy and depth of the embeddings by providing crucial prior knowledge. Besides, it would reduce vocabulary sizes, thus optimizing model performance and efficiency. In conclusion, our study confirms the potential of language models in healthcare data analysis, particularly in understanding patient trajectories in intensive care.

## Funding statement

This work was funded by the Innosuisse – Schweizerische Agentur für Innovationsförderung – project no.: 55441.1 IP ICT.

## Data Availability

The MIMIC-IV dataset used in this study is publicly available upon request from the PhysioNet repository (https://physionet.org/content/mimiciv/2.1/). The exact queries and scripts used for data extraction and analysis from this dataset are available at https://github.com/ds4dh/medical_concept_representation.

## Footnotes

1 https://github.com/ds4dh/medical_concept_representation

2 https://github.com/ds4dh/medical_concept_representation/tree/main/data/datasets/mimic-iv-2.2/maps

3 https://icd.who.int/browse10/2010/en

## Notes

### Competing Interest Statement

The authors have declared no competing interest.

### Author Declarations

Data used in this study were sourced exclusively from the MIMIC-IV database, a publicly available, de-identified human dataset hosted on the PhysioNet repository (https://physionet.org/content/mimiciv/2.2/). Access to the MIMIC-IV database required completion of mandatory coursework on data privacy and human subjects research protection. The creation and use of the MIMIC-IV database have been approved by the Institutional Review Boards of Beth Israel Deaconess Medical Center (Boston, MA) and the Massachusetts Institute of Technology (Cambridge, MA). This study, involving secondary analysis of pre-existing, de-identified data, does not classify as human subjects research, and therefore, did not necessitate additional ethical approval.

### Summary of Updates

This revised version enhances the clarity, robustness, and interpretability of our study. We eliminated the subsampling of patients displayed in Figure 7, and instead computed t-SNE over the entire test dataset. Moreover, we shifted our analysis approach from direct embedding comparisons to using Representational Dissimilarity Matrices (RDM). We enhanced the statistical robustness of our results by incorporating bootstrapping to estimate the variability of model prediction performances and include confidence intervals in Figures 6, 8, and 9. We also updated our interpretation of the results in the discussion. We refined the introduction to provide a clearer overview of related work, and revised the methods section to include more precise details about our experimental setup.

