## Appendix A1 for "Comparing neural language models for medical concept representation and patient trajectory prediction"

Below, we provide a detailed description of each token category used to build the patient trajectories from the MIMIC dataset (see section 2.1).

Using the MIMIC table “admissions”, we associated length-of-stay, readmission, and mortality outcomes to each patient trajectory. These tokens were then used as labels for length-of-stay, readmission, and mortality prediction tasks (more details in subsection 2.4.2), and defined as follows. A patient trajectory was considered length-of-stay-positive, i.e., extended status, if the time elapsed between admission time and discharge time was longer than 7 days, and length-of-stay-negative, i.e., normal status, otherwise. A patient trajectory was considered readmission-positive, i.e., readmitted status, if the associated patient was readmitted less than 30 days after discharge time, and readmission-negative, i.e., no-readmission status, otherwise. A patient trajectory was considered mortality-positive, i.e., dead status, if the associated patient passed away during the stay or less than 30 days after discharge time, and mortality-negative, i.e., alive status, otherwise. All outcome tokens started with the suffix “lbl_”.

Patient demographic information was extracted from the MIMIC tables “patient” and “admissions”, and included gender, age and race. To express the age of a patient as a token, we used 4 categories: “young” for age < 21 yo.; “adult” for age >= 21 yo. and < 50 yo.; “middle” for age >= 50 yo. and < 65 yo.; and “old” for age >= 65 yo. We used a custom dictionary to map races included in the dataset to 33 different race labels (including “unknown”; see Table S1, Suppl. Inf.). Most races were expressed as a set of two tokens (e.g., “latino-mexico”). All demographic tokens started with the suffix “dem_”.

Administrative events were extracted from the table “transfers” and mapped to local terminologies, such as for hospital locations (e.g., PACU for Post-Anesthesia Care Unit). Everytime a patient is transferred to a new care unit, the information is stored and linked to an entrance time, which we used to sort location tokens within patient trajectories. We used a custom dictionary (see Table S2, Suppl. Inf.) to map any location included in the dataset to 40 different location labels (including “unknown”). Some hospital locations were expressed with several tokens (up to 3, e.g., “med-surg-icu”). All location tokens started with the suffix “loc_”.

Medical events were mapped either to standard biomedical terminologies (e.g., ICD10 for diagnosis and procedure codes, ATC for medications) or to local terminologies (for laboratory test results). ICD10-CM codes for diagnoses and reason for visits were extracted from the table “diagnoses_icd” and ICD10-PCS codes for procedure events were extracted from the table “procedures_icd”. We also considered ICD9 codes appearing in the tables and converted them to their ICD10 equivalents using the mapping available at the National Bureau of Economics^[1]^. Diagnoses do not have associated timestamps and were hence added early in the sequence as patients tend to have a main diagnosis before entering the ICU. Still, these tokens were sorted by level of importance (“seq_num” field in the MIMIC-IV database). Procedure events are associated with a chart date when recorded in the database, which we used to sort procedure tokens within patient trajectory sequences. Medication prescription events were extracted from the table “prescriptions”. Everytime a medication is prescribed to a patient, it is associated with a prescribed start time, which we used to sort the medication tokens within patient trajectory sequences. Medications are stored in MIMIC-IV as Generic Sequence Number (GSN) or National Drug Code (NDC) entries. We converted any of these codes to its Anatomical Therapeutic Chemical (ATC) code equivalent. ATC codes are hierarchically arranged following the target anatomical system of medications, as well as their therapeutic, pharmacological, and chemical effects. To ensure most medication entries were correctly converted to ATC codes, we built a custom mapping table as follows. First, we built an NDC to ATC mapping, using the work of Kury et al. (2017)^[2]^ that queries the online RxNorm API^[3]^ in an automated way. Then, we built a GSN to ATC mapping, combining the table available at the NCBO BioPortal^[4]^, which maps ATC codes to different drug names, and the MIMIC-IV table “medrecon”, which does the same for GSN codes. Finally, we combined the GSN to ATC and NDC to ATC mappings into a single table, and manually searched for about 1,300 missing ATC code mappings, verifying the retrieved information with the WHO-ATC indexing website^[5]^. We make the final table available at our repository^[6]^. Laboratory measurement events were extracted from the MIMIC table “labevents”. Every lab test in MIMIC-IV comes with the time at which it was acquired, which we used to sort lab event tokens within patient trajectories. To reduce the number of tokens in each patient trajectory and increase the relevance of lab events, lab event tokens were included only if they indicated an abnormal result, i.e., a measurement that is outside the referenced normal ranges. To construct lab event tokens, we simply used the ids that are assigned to any laboratory concept in MIMIC-IV. These ids do not carry any semantic meaning.

**Table S1. Dictionary used to map races included in MIMIC-IV to race labels used to build patient trajectory sequences.** Races were extracted from the field “race” of the table “admissions” of the MIMIC-IV dataset.

| **Race (field “race” of MIMIC-IV table “admissions”)** | **Race label** |
| --- | --- |
| BLACK/AFRICAN AMERICAN | BLACK-USA |
| HISPANIC/LATINO - COLUMBIAN | LATINO-CLMB |
| HISPANIC/LATINO - CUBAN | LATINO-CUBA |
| BLACK/CAPE VERDEAN | BLACK-CAPV |
| WHITE - RUSSIAN | WHITE-RUSS |
| BLACK/AFRICAN | BLACK-AFRIC |
| ASIAN | ASIAN |
| BLACK/CARIBBEAN ISLAND | BLACK-CARB |
| ASIAN - KOREAN | ASIAN-KOR |
| MULTIPLE RACE/ETHNICITY | MULTIPLE |
| OTHER | UNKNOWN |
| HISPANIC/LATINO - PUERTO RICAN | LATINO-PRICO |
| ASIAN - ASIAN INDIAN | ASIAN-INDIAN |
| HISPANIC/LATINO - MEXICAN | LATINO-MXCO |
| WHITE - BRAZILIAN | WHITE-BRZL |
| HISPANIC/LATINO - HONDURAN | LATINO-HOND |
| WHITE - EASTERN EUROPEAN | WHITE-EAST |
| HISPANIC/LATINO - CENTRAL AMERICAN | LATINO-CTRA |
| ASIAN - SOUTH EAST ASIAN | ASIAN-SOUTH |
| AMERICAN INDIAN/ALASKA NATIVE | NATIVE-USA |
| HISPANIC/LATINO - SALVADORAN | LATINO-SLVD |
| ASIAN - CHINESE | ASIAN-CHINA |
| NATIVE HAWAIIAN OR OTHER PACIFIC ISLANDER | NATIVE-HAWAI |
| HISPANIC/LATINO - GUATEMALAN | LATINO-GUAT |
| HISPANIC OR LATINO | LATINO-HISP |
| UNABLE TO OBTAIN | UNKNOWN |
| SOUTH AMERICAN | LATINO |
| PATIENT DECLINED TO ANSWER | UNKNOWN |
| PORTUGUESE | WHITE-PORT |
| WHITE | WHITE |
| HISPANIC/LATINO - DOMINICAN | LATINO-DOMI |
| UNKNOWN | UNKNOWN |
| WHITE - OTHER EUROPEAN | WHITE-OTHER |

**Table S2. Dictionary used to map locations included in MIMIC-IV to location labels used to build patient trajectory sequences.** Locations were extracted from the field “careunit” of the table “transfers” of the MIMIC-IV dataset. Note that the “nan” value was used to retrieve discharge events, since there is a one-to-one mapping between “nan” values in the “careunit” field and “discharge” value in the “eventtype” field of the table “transfers”.

| **Location (field “careunit” of MIMIC-IV table “transfers”)** | **Location label** |
| --- | --- |
| Unknown | UNKNOWN |
| Med/Surg/GYN | MED-SURG-GYN |
| Surgical Intensive Care Unit (SICU) | SURG-ICU |
| Neuro Stepdown | NEURO-STEP |
| Neurology | NEURO |
| Coronary Care Unit (CCU) | CORO-CU |
| Medicine/Cardiology | MED-CARD |
| Medical/Surgical Intensive Care Unit (MICU/SICU) | MED-SURG-ICU |
| Cardiology Surgery Intermediate | CARD-SURG-INTER |
| Obstetrics Postpartum | PREG-POST |
| PACU | ANEST-POST-CU |
| Observation | OBS |
| Medical Intensive Care Unit (MICU) | MED-ICU |
| nan | DISCH |
| Medicine/Cardiology Intermediate | MED-CARD-INTER |
| Emergency Department | ED |
| Cardiac Vascular Intensive Care Unit (CVICU) | CARD-VASC-CU |
| Vascular | VASC |
| Hematology/Oncology | HEMA-ONCO |
| Neuro Intermediate | NEURO-INTER |
| Med/Surg | MED-SURG |
| Surgery | SURG |
| Medicine | MED |
| Hematology/Oncology Intermediate | HEMA-ONCO-INTER |
| Psychiatry | PSYCH |
| Emergency Department Observation | OBS-ED |
| Obstetrics (Postpartum & Antepartum) | PREG |
| Surgery/Pancreatic/Biliary/Bariatric | SURG-PANCR |
| Neuro Surgical Intensive Care Unit (Neuro SICU) | NEURO-SURG-ICU |
| Medical/Surgical (Gynecology) | MED-SURG-GYN |
| Discharge Lounge | DISCH-LNGE |
| Labor & Delivery | PREG-LABOR |
| Med/Surg/Trauma | MED-SURG-TRAUMA |
| Cardiac Surgery | SURG-CARD |
| Transplant | TRANS |
| Thoracic Surgery | SURG-THORAX |
| Surgery/Trauma | SURG-TRAUMA |
| Cardiology | CARD |
| Trauma SICU (TSICU) | SURG-TRAUMA-ICU |
| Obstetrics Antepartum | PREG-ANTE |

1. https://www.nber.org/research/data/icd-9-cm-and-icd-10-cm-and-icd-10-pcs-crosswalk-or-general-equivalence-mappings
2. Kury FS, Bodenreider O (2017) Mapping US FDA National Drug Codes to Anatomical-Therapeutic-Chemical Classes using RxNorm. AMIA
3. Nelson SJ, Zeng K, Kilbourne J, Powell T, Moore R (2011) Normalized names for clinical drugs: RxNorm at 6 years. Journal of the American Medical Informatics Association 18:441–448
4. https://bioportal.bioontology.org/ontologies/ATC
5. https://www.whocc.no/atc_ddd_index/
6. https://github.com/ds4dh/medical_concept_representation/tree/main/data/datasets/mimic-iv-2.2/maps
