## Appendix A2 for "Comparing neural language models for medical concept representation and patient trajectory prediction"

| **Category** | **Subcategories** | **Label** | **Remark** |
| --- | --- | --- | --- |
| **ICD10-CM** | ‘A’, ‘B’ | Certain infectious and parasitic diseases | <1% |
|  | 'C', 'D0', 'D1', 'D2', 'D3', 'D4' | Neoplasms |  |
|  | 'D5', 'D6', 'D7', 'D8', 'D9' | Diseases of the blood and blood-forming organs and certain disorders involving the immune mechanism |  |
|  | ‘E’ | Endocrine, nutritional and metabolic diseases | <1% |
|  | ‘F’ | Mental, Behavioral and Neurodevelopmental disorders |  |
|  | ‘G’ | Diseases of the nervous system |  |
|  | 'H0', 'H1', 'H2', 'H3', 'H4', 'H5' | Diseases of the eye and adnexa |  |
|  | 'H6', 'H7', 'H8', 'H9' | Diseases of the ear and mastoid process |  |
|  | ‘I’ | Diseases of the circulatory system |  |
|  | ‘J’ | Diseases of the respiratory system |  |
|  | ‘K’ | Diseases of the digestive system |  |
|  | ‘L’ | Diseases of the skin and subcutaneous tissue |  |
|  | ‘M’ | Diseases of the musculoskeletal system and connective tissue |  |
|  | ‘N’ | Diseases of the genitourinary system |  |
|  | ‘O’ | Pregnancy, childbirth and the puerperium |  |
|  | ‘P’ | Certain conditions originating in the perinatal period | <1% |
|  | ‘Q’ | Congenital malformations, deformations and chromosomal abnormalities | <1% |
|  | ‘R’ | Symptoms, signs and abnormal clinical and laboratory findings, not elsewhere classified | Not visualized |
|  | ‘S’, ‘T’ | Injury, poisoning and certain other consequences of external causes | Separated into ‘S’ and ‘T’ |
|  | ‘U’ | Codes for special purposes | <1% |
|  | ‘V’, ‘W’, ‘X’, ‘Y’ | External causes of morbidity | <1% |
|  | ‘Z’ | Factors influencing health status and contact with health services |  |
| **ICD10-PCS** | ‘00’ | Medical and Surgical - Central Nervous System and Cranial Nerves |  |
|  | ‘01’ | Medical and Surgical - Peripheral Nervous System | <1% |
|  | ‘02’ | Medical and Surgical - Heart and Great Vessels | <1% |
|  | ‘03’ | Medical and Surgical - Upper Arteries | <1% |
|  | ‘04’ | Medical and Surgical - Lower Arteries |  |
|  | ‘05’ | Medical and Surgical - Upper Veins | <1% |
|  | ‘06’ | Medical and Surgical - Lower Veins | <1% |
|  | ‘07’ | Medical and Surgical - Lymphatic and Hemic Systems | <1% |
|  | ‘08’ | Medical and Surgical - Eye | <1% |
|  | ‘09’ | Medical and Surgical - Ear, Nose, Sinus | <1% |
|  | ‘0B’ | Medical and Surgical - Respiratory System |  |
|  | ‘0C’ | Medical and Surgical - Mouth and Throat | <1% |
|  | ‘0D’ | Medical and Surgical - Gastrointestinal System |  |
|  | ‘0F’ | Medical and Surgical - Hepatobiliary System and Pancreas |  |
|  | ‘0G’ | Medical and Surgical - Endocrine System | <1% |
|  | ‘0H’ | Medical and Surgical - Skin and Breast |  |
|  | ‘0J’ | Medical and Surgical - Subcutaneous Tissue and Fascia |  |
|  | ‘0K’ | Medical and Surgical - Muscles | <1% |
|  | ‘0L’ | Medical and Surgical - Tendons | <1% |
|  | ‘0M’ | Medical and Surgical - Bursae and Ligaments | <1% |
|  | ‘0N’ | Medical and Surgical - Head and Facial Bones | <1% |
|  | ‘0P’ | Medical and Surgical - Upper Bones | <1% |
|  | ‘0Q’ | Medical and Surgical - Lower Bones |  |
|  | ‘0R’ | Medical and Surgical - Upper Joints | <1% |
|  | ‘0S’ | Medical and Surgical - Lower Joints |  |
|  | ‘0T’ | Medical and Surgical - Urinary System |  |
|  | ‘0U’ | Medical and Surgical - Female Reproductive System |  |
|  | ‘0V’ | Medical and Surgical - Male Reproductive System | <1% |
|  | ‘0W’ | Medical and Surgical - Anatomical Regions, General |  |
|  | ‘0X’ | Medical and Surgical - Anatomical Regions, Upper Extremities | <1% |
|  | ‘0Y’ | Medical and Surgical - Anatomical Regions, Lower Extremities | <1% |
|  | ‘1’ | Obstetrics | <1% |
|  | ‘2’ | Placement | <1% |
|  | ‘3’ | Administration |  |
|  | ‘4’ | Measurement and Monitoring | <1% |
|  | ‘5’ | Extracorporeal or Systemic Assistance and Performance | <1% |
|  | ‘6’ | Extracorporeal or Systemic Therapies | <1% |
|  | ‘7’ | Osteopathic | <1% |
|  | ‘8’ | Other Procedures | <1% |
|  | ‘9’ | Chiropractic | <1% |
|  | ‘B’ | Imaging |  |
|  | ‘C’ | Nuclear Medicine | <1% |
|  | ‘D’ | Radiation Therapy | <1% |
|  | ‘F’ | Physical Rehabilitation and Diagnostic Audiology | <1% |
|  | ‘G’ | Mental Health | <1% |
|  | ‘H’ | Substance Abuse Treatment | <1% |
|  | ‘X’ | New Technology | <1% |
| **ATC codes** | ‘A’ | Alimentary Tract and Metabolism |  |
|  | ‘B’ | Blood and Blood Forming Organs |  |
|  | ‘C’ | Cardiovascular System |  |
|  | ‘D’ | Dermatologicals |  |
|  | ‘G’ | Genito-Urinary System and Sex Hormones |  |
|  | ‘H’ | Systemic Hormonal Preparations, Excl. Sex Hormones and Insulins | <1% |
|  | ‘J’ | Anti Infectives for Systemic Use |  |
|  | ‘L’ | Antineoplastic and Immunomodulating Agents |  |
|  | ‘M’ | Musculo-Skeletal System | <1% |
|  | ‘N’ | Nervous System |  |
|  | ‘P’ | Antiparasitic Products, Insecticides and Repellents | <1% |
|  | ‘R’ | Respiratory System |  |
|  | ‘S’ | Sensory Organs |  |
|  | ‘V’ | Various | <1% |

**Table S3. Detail of the subcategories used for the visualization in Figure 4.** Note that we only kept subcategories that, on the one hand, held at least 1% of the codes in the terminology and, on the other hand, represented at least 1% of the tokens of the training dataset.
