## Appendix A4 for "Comparing neural language models for medical concept representation and patient trajectory prediction"

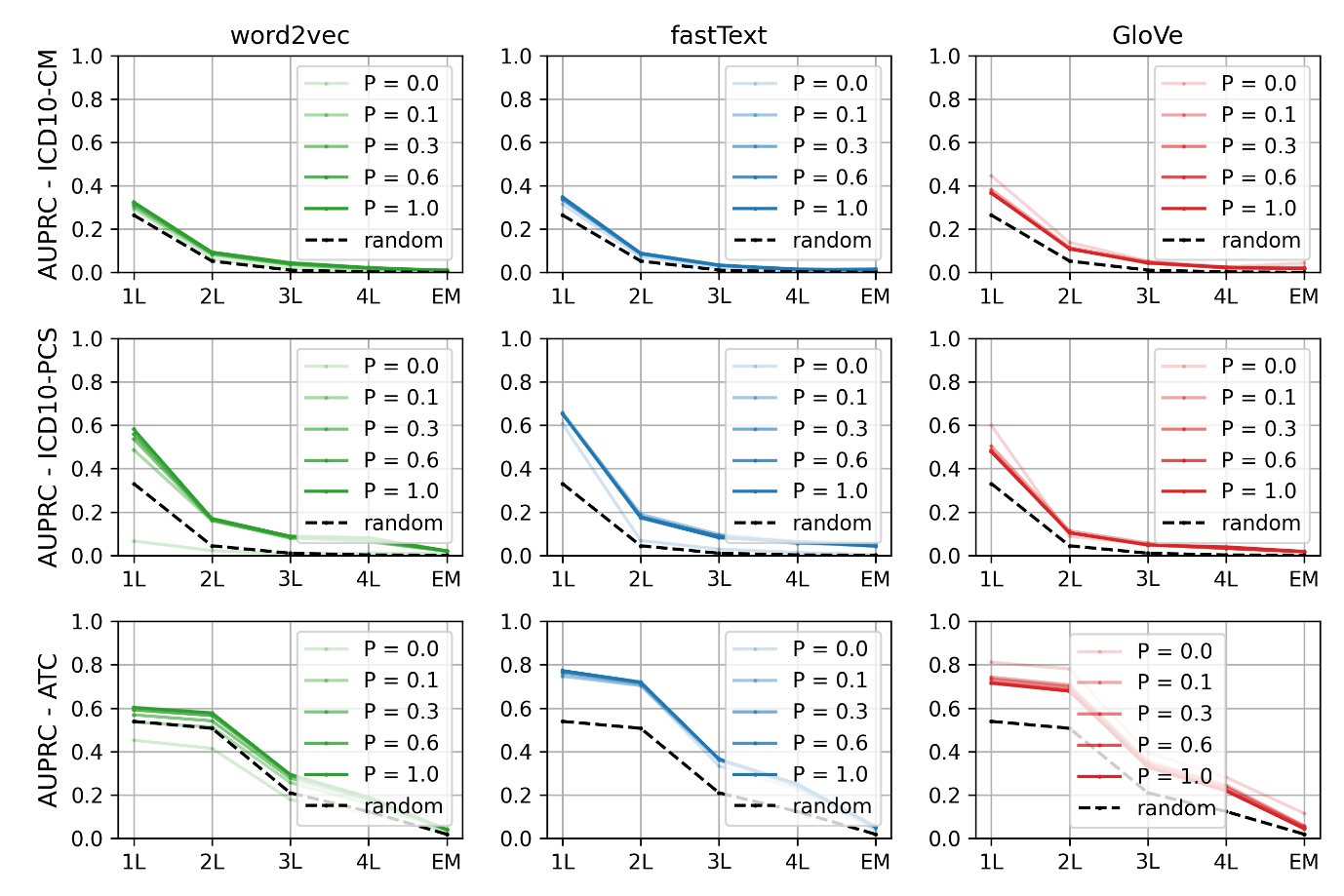

**Figure S2. AUPRC obtained with word2vec, fastText and GloVe for medical concept prediction tasks (see Figure 8).** L stands for the lenient letter match. For example, 1L means that precision and recall were computed based on the first letter of the codes only. EM means that an exact match was required between a model prediction and a target code. P stands for the different proportions of the patient trajectories given as input to the models (see section 2.4). The reported means were calculated using the Bag of Little Bootstraps algorithm with n_bootstraps = 100, n_subsamples = 100, subsample_size = 10,000, and standard deviations were not added for readability.
