## Appendix A6 for "Comparing neural language models for medical concept representation and patient trajectory prediction"

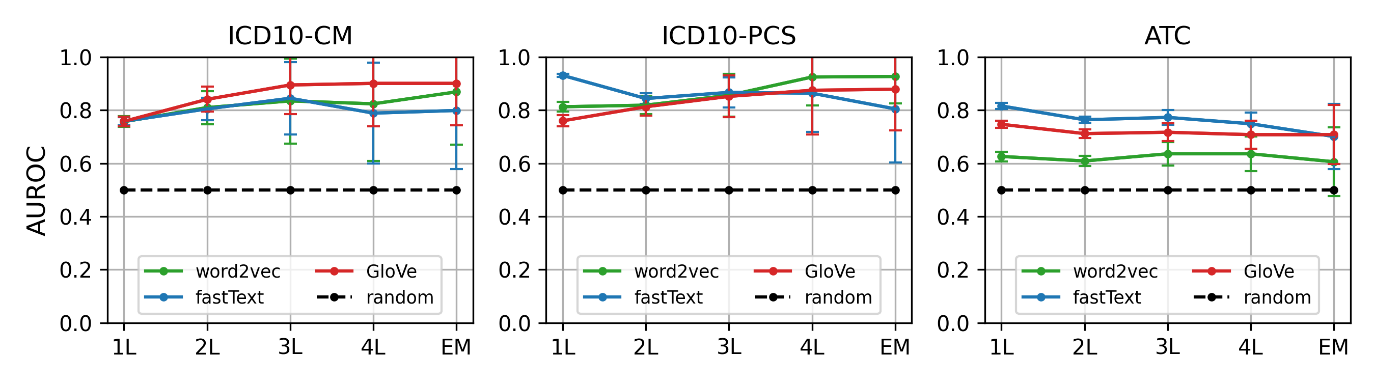

**Figure S4. AUROC obtained with word2vec, fastText and GloVe for the same prediction tasks as presented in Figure 9.** We used model embeddings to predict the most important ICD10-CM code of the patient trajectory, as well as the next ICD10-PCS and ATC code, given the medical events that happened up to the target token. L stands for the lenient letter match. For example, 1L means that precision and recall were computed based on the first letter of the codes only. EM means that an exact match was required between a model prediction and a target code. The reported means were calculated using the Bag of Little Bootstraps algorithm with n_bootstraps = 100, n_subsamples = 100, subsample_size = 10,000, and standard deviations were not added for readability.
